# Intellectual enrichment and genetic modifiers of cognitive function in Huntington’s disease

**DOI:** 10.1101/2022.04.09.22273637

**Authors:** Marina Papoutsi, Michael Flower, Davina Hensman, Peter Holmans, Carlos Estevez-Fraga, Eileanor Johnson, Rachael I. Scahill, Geraint Rees, Douglas Langbehn, Sarah J. Tabrizi, the Track-HD Investigators

**Author notes:** Corresponding author: Dr Marina Papoutsi, Address: UCL Huntington’s disease centre, Russell Square House, 10-12 Russell Square, London, WC1B 5EH, UK.

## Abstract

An important step towards the development of treatments for cognitive impairment in ageing and neurodegenerative diseases is to identify genetic and environmental modifiers of cognitive function and understand the mechanism by which they exert an effect. In Huntington’s disease, the most common autosomal dominant dementia, a small number of studies have identified intellectual enrichment, i.e. a cognitively stimulating lifestyle, and genetic polymorphisms as potential modifiers of cognitive function. The aim of our study was to further investigate the relationship and interaction between genetic factors and intellectual enrichment on cognitive function and brain atrophy in Huntington’s disease. For this purpose, we analysed data from Track-HD, a multi-centre longitudinal study in Huntington’s disease gene-carriers, and focused on the role of intellectual enrichment (estimated at baseline) and the genes *FAN1, MSH3, BDNF, COMT* and *MAPT* in predicting cognitive decline and brain atrophy. We found that carrying the 3a allele in the *MSH3* gene had a positive effect on global cognitive function and brain atrophy in multiple cortical regions, such that 3a allele carriers had a slower rate of cognitive decline and atrophy compared to non-carriers, in agreement with its role in somatic expansion instability. No other genetic predictor had a significant effect on cognitive function and the effect of *MSH3* was independent of intellectual enrichment. Intellectual enrichment also had a positive effect on cognitive function; participants with higher intellectual enrichment, ie. those who were better educated, had higher verbal IQ and performed an occupation that was intellectually engaging, had better cognitive function overall, in agreement with previous studies in Huntington’s disease and other dementias. We also found that intellectual enrichment interacted with the *BDNF* gene, such that the positive effect of intellectual enrichment was greater in Met66 allele carriers than non-carriers. A similar relationship was also identified for changes in whole brain and caudate volume; the positive effect of intellectual enrichment was greater for Met66 allele carriers, rather than non-carriers. In summary, our study provides additional evidence for the beneficial role of intellectual enrichment and carrying the 3a allele in *MSH3* in cognitive function in Huntington’s disease and their mechanism of action.

## Introduction

Huntington’s disease is a genetic, neurodegenerative disorder caused by an abnormal CAG repeat expansion in the *HTT* gene. It is characterised by a triad of symptoms, motor, psychiatric and cognitive. All Huntington’s disease gene-carriers will eventually develop dementia^1^, but there is substantial variability in its onset and severity, which cannot be explained fully by CAG repeat length and age. Cognitive impairment is present in Huntington’s disease gene-carriers many years before predicted disease onset and in the absence of motor symptoms^2^. However, research on the genetic and environmental factors that contribute to this variability in cognitive impairment in Huntington’s disease is still limited.

Individual differences in cognitive function and rate of decline have been studied extensively in ageing and Alzheimer’s disease. One prominent hypothesis is that of brain maintenance^3^, according to which the primary determinant of preserved cognitive function is lower levels of pathology and slower rate of neurodegeneration. However, it has also been observed that individual differences in cognitive impairment exist despite similar levels of neurodegeneration, which led to the theory of cognitive reserve^4^. Although the genetic and environmental factors that support brain maintenance and cognitive reserve in ageing and dementia are not all known, lifelong participation in intellectual activities, also known as intellectual enrichment^5^, as well as genetic polymorphisms^6^, have been associated with preserved cognitive function and mechanisms of brain maintenance and cognitive reserve. Genetic factors and intellectual enrichment have also been shown to interact and enhance their effect on brain structure and cognition^7–9^.

In Huntington’s disease, a small number of studies have so far examined the role of genetic polymorphisms and lifestyle factors on individual differences in cognitive function. More specifically, education and participation in lifelong intellectual activities^10–12^, as well as a number of genes, including *FAN1*^13^, *COMT*^14^*, MSH3*^15^ and *MAPT*^16^, predict cognitive function. Two of these studies have also provided preliminary evidence that intellectual enrichment is associated with less striatal atrophy^11, 12^, suggesting that it supports greater brain maintenance.

The aim of our work was to provide evidence regarding the effect of intellectual enrichment and genetic factors on cognitive function and brain structure in Huntington’s disease. For this purpose, we retrospectively analysed data from a multi-centre, longitudinal study, Track-HD^17–20^, that measured change in behaviour and brain structure over 3-years in individuals with the Huntington’s disease gene mutation in pre-manifest (maximum 15 years from predicted onset) and early stages of the disease. We quantified lifetime intellectual enrichment using level of education, premorbid IQ and occupational cognitive demands^11^ measured at baseline. In terms of genetic polymorphisms, we selected common polymorphisms that have been previously associated with cognitive function in Huntington’s disease (*COMT*, *BDNF, FAN1, MSH3* and *MAPT*).

## Materials and Methods

### Participants

Track-HD is a multi-centre, 4-year observational study in Huntington’s disease gene-carriers and matched controls. A full description of the Track-HD study has been previously reported^17–20^. In summary, 243 Huntington’s disease gene-carriers (both manifest and pre-manifest) and 123 matched controls were recruited across four sites (London, UK; Paris, France; Leiden, The Netherlands, and Vancouver, Canada). Local ethics committees approved the study at each site and all participants provided written informed consent according to the Declaration of Helsinki.

The Track-HD study included detailed measures of brain structure, cognitive and motor function, in addition to information regarding education, pre-morbid IQ and profession. Blood for DNA analyses was also collected. Table 1 shows details of the measures that were used in this study and the number of participants included (split by visit for longitudinal measures). Data from all Huntington’s disease gene-carriers with at least one follow-up visit (n = 229), irrespective of disease diagnosis, were used for the analyses. Data from the matched control group were only used to create standardized scores of cognitive performance in the gene-carrier group (demographic information on the control group is provided in Supplementary Table 1).

**Table 1:**
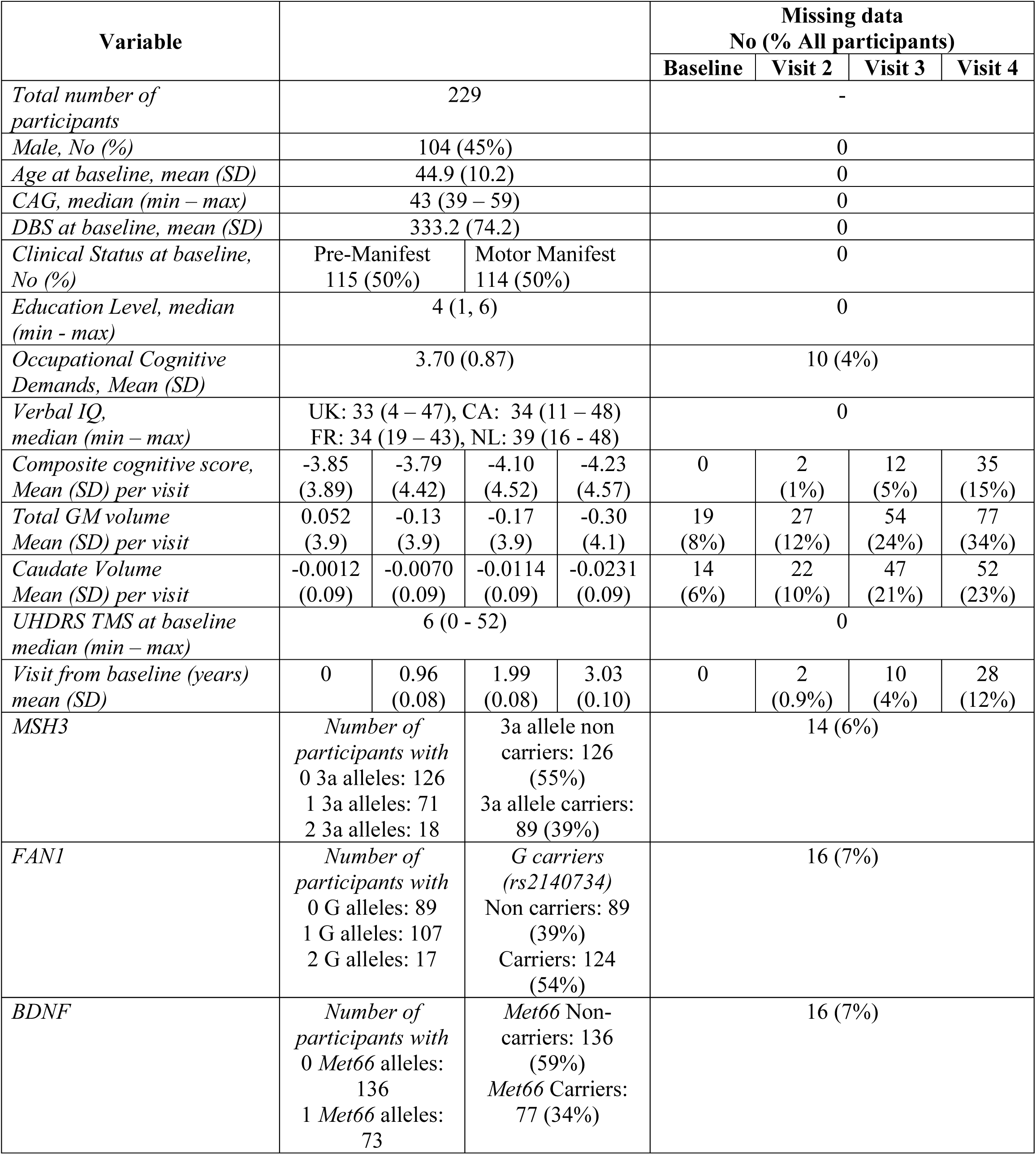
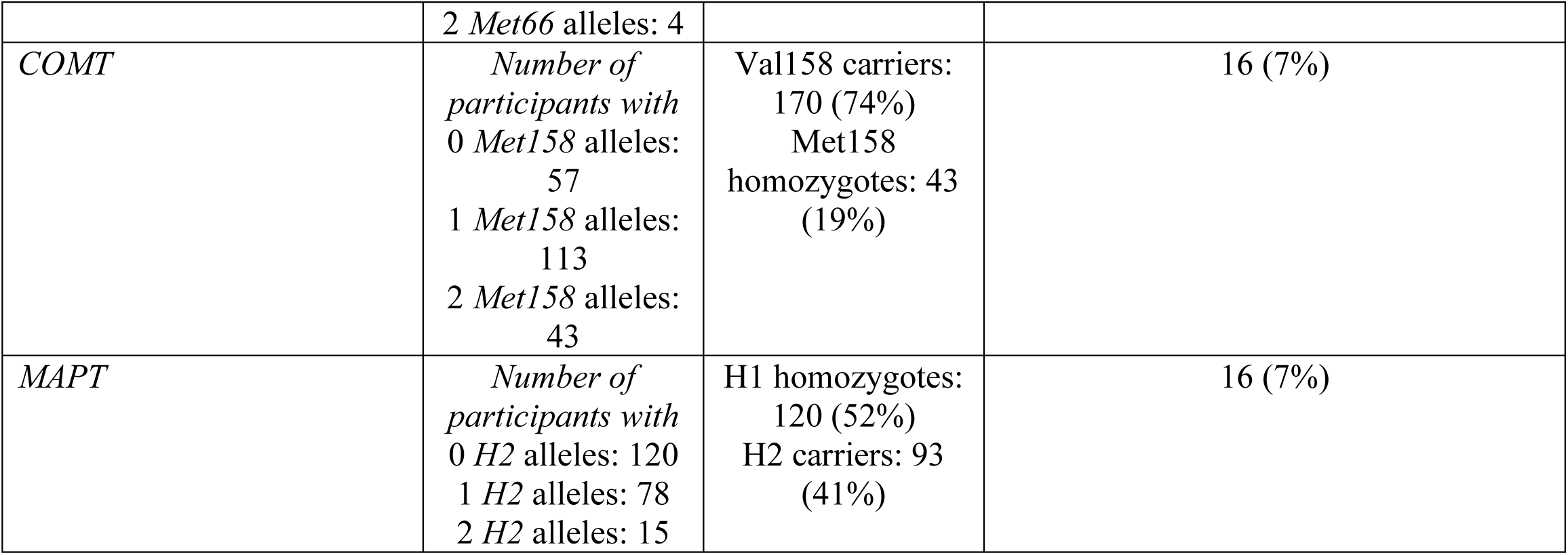
HD gene-carrier demographics and clinical characteristics

| Variable |  |  |  |  | Missing data<br>No (% All participants) |  |  |  |
| --- | --- | --- | --- | --- | --- | --- | --- | --- |
|  |  |  |  |  | Baseline | Visit 2 | Visit 3 | Visit 4 |
| Total number of participants | 229 |  |  |  | - |  |  |  |
| Male, No (%) | 104 (45%) |  |  |  | 0 |  |  |  |
| Age at baseline, mean (SD) | 44.9 (10.2) |  |  |  | 0 |  |  |  |
| CAG, median (min – max) | 43 (39 – 59) |  |  |  | 0 |  |  |  |
| DBS at baseline, mean (SD) | 333.2 (74.2) |  |  |  | 0 |  |  |  |
| Clinical Status at baseline, No (%) | Pre-Manifest<br>115 (50%) |  | Motor Manifest<br>114 (50%) |  | 0 |  |  |  |
| Education Level, median (min - max) | 4 (1, 6) |  |  |  | 0 |  |  |  |
| Occupational Cognitive Demands, Mean (SD) | 3.70 (0.87) |  |  |  | 10 (4%) |  |  |  |
| Verbal IQ, median (min – max) | UK: 33 (4 – 47), CA: 34 (11 – 48)<br>FR: 34 (19 – 43), NL: 39 (16 - 48) |  |  |  | 0 |  |  |  |
| Composite cognitive score, Mean (SD) per visit | -3.85<br>(3.89) | -3.79<br>(4.42) | -4.10<br>(4.52) | -4.23<br>(4.57) | 0 | 2<br>(1%) | 12<br>(5%) | 35<br>(15%) |
| Total GM volume Mean (SD) per visit | 0.052<br>(3.9) | -0.13<br>(3.9) | -0.17<br>(3.9) | -0.30<br>(4.1) | 19<br>(8%) | 27<br>(12%) | 54<br>(24%) | 77<br>(34%) |
| Caudate Volume Mean (SD) per visit | -0.0012<br>(0.09) | -0.0070<br>(0.09) | -0.0114<br>(0.09) | -0.0231<br>(0.09) | 14<br>(6%) | 22<br>(10%) | 47<br>(21%) | 52<br>(23%) |
| UHDRS TMS at baseline median (min – max) | 6 (0 - 52) |  |  |  | 0 |  |  |  |
| Visit from baseline (years) mean (SD) | 0 | 0.96<br>(0.08) | 1.99<br>(0.08) | 3.03<br>(0.10) | 0 | 2<br>(0.9%) | 10<br>(4%) | 28<br>(12%) |
| MSH3 | Number of participants with<br>0 3a alleles: 126<br>1 3a alleles: 71<br>2 3a alleles: 18 |  | 3a allele non carriers: 126 (55%)<br>3a allele carriers: 89 (39%) |  | 14 (6%) |  |  |  |
| FAN1 | Number of participants with<br>0 G alleles: 89<br>1 G alleles: 107<br>2 G alleles: 17 |  | G carriers (rs2140734)<br>Non carriers: 89 (39%)<br>Carriers: 124 (54%) |  | 16 (7%) |  |  |  |
| BDNF | Number of participants with<br>0 Met66 alleles: 136<br>1 Met66 alleles: 73 |  | Met66 Non-carriers: 136 (59%)<br>Met66 Carriers: 77 (34%) |  | 16 (7%) |  |  |  |

|  |  |  |  |
| --- | --- | --- | --- |
|  | 2 *Met66* alleles: 4 |  |  |
| *COMT* | *Number of participants with*<br>0 *Met158* alleles: 57<br>1 *Met158* alleles: 113<br>2 *Met158* alleles: 43 | Val158 carriers: 170 (74%)<br>Met158 homozygotes: 43 (19%) | 16 (7%) |
| *MAPT* | *Number of participants with*<br>0 *H2* alleles: 120<br>1 *H2* alleles: 78<br>2 *H2* alleles: 15 | H1 homozygotes: 120 (52%)<br>H2 carriers: 93 (41%) | 16 (7%) |

### Global Cognitive Function

To quantify cognitive function and change over three years we created a composite score from the available cognitive measures. This composite score represents global cognitive function and is comprised of the following measures from the Track-HD cognitive battery^17^: number correct in 90s from the Symbol Digit Modality Test (SDMT) (a measure of processing speed), number correct in 45s from the Stroop word reading test (a measure of psychomotor speed), number correct adjusted for guessing (greater than 0 means better than chance) for the five items condition from the Spot the Change task (a measure of working memory), number correct for negative emotions from the Emotion Recognition task (a measure of facial emotion recognition), and variability in inter-tap interval in a paced tapping task at 3Hz (a measure of temporal precision). These tasks were included in all four visits, in addition to the Circle Tracing task. However, that task was excluded from the composite score because of large practice effects that persisted across all visits^21^. The remaining five measures were then used to create a composite score of global cognitive function.

To calculate the composite score, raw values were transformed to z-scores using the mean and standard deviation of the control group at baseline and then summed. In the paced tapping task, the reciprocal of the variance in inter-tap interval was used, such that a higher value indicated better performance (i.e lower variability), consistent with all other measures. Therefore, higher values in the composite score indicate better performance. When computing the composite score, if one or two measures were missing, they were replaced by the mean z-score of the existing measures. This was the case for 21 participants in visit one, seven participants in visit two, three participants in visit three, and 16 participants in visit four respectively. If a participant had less than three out of five measures available for a visit, then we did not compute a composite score for that visit and therefore this participant-visit was not included in the analyses with cognitive function as a variable.

### Genetic Polymorphisms

Track-HD subjects were genotyped using Illumina Omni2.5v1.1 arrays^22^ and genotypes were extracted using PLINK^23^. Because the main outcome measure was cognitive function, we focused on SNPs that had been previously associated with cognitive and psychomotor function in Huntington’s disease. Based on previous literature we therefore selected the following SNPs: rs4680 on chromosome 22^14^ in *COMT*, which relates to dopamine metabolism, rs9468 on chromosome 17^16^ in *MAPT*, which relates to tau protein production, rs2140734 on chromosome 15 near *FAN1*^13, 24^, which is involved in DNA repair, and a polymorphic repeat expansion in exon 1 on chromosome 5 of *MSH3* ^15, 22, 25, 26^, which is involved in DNA mismatch repair. In addition, we tested another polymorphism, rs6265 (Val66Met) on chromosome 11 in *BDNF*, which encodes the Val66Met polymorphism and regulates BDNF expression. Although the role of the Val66Met polymorphism in Huntington’s disease remains unknown, it has been consistently associated with cognitive function in ageing and dementia^6, 27–29^ and *BNDF* expression maybe affected by Huntington’s disease pathology ^30, 31^. It also interacts with intellectual enrichment factors to predict cognitive function^7, 32^: it was therefore relevant to our research question.

A previous study showed that the gene *TREM2* (rs75932628) has a role in cognitive function in Huntington’s disease^33^. However, the minor allele frequency is very low in the population (0.005% in Europeans in 1000 Genomes project data phase 3) and therefore we did not include it in our analyses.

All genetic predictors were coded as having a binary, dominant effect, similar to the approach of Vuono et al.^16^. Supplementary Table 2 shows how each binary predictor was coded. In more detail, the *MSH3* predictor was coded for the presence of the three-repeat allele (3a)^25^. The rs2140734 (*FAN1*) predictor was coded for the presence of the minor allele G. It is also important to highlight that this SNP is in complete linkage disequilibrium with minor allele C in rs3512. The latter has been more widely examined in other studies and shown to be associated with age of onset and disease progression in Huntington’s disease^22, 24, 34^. The *MAPT* predictor distinguished between H1 haplotype homozygotes and H2 carriers. Carrying the minor allele C in rs9468 tags for the H2 haplotype, whereas carrying the allele T tags for the H1 haplotype^35^. The *COMT* (rs4680) predictor variable distinguished Met158 homozygotes from carriers of the Val158 allele^14^. The *BDNF* (rs6265) predictor variable distinguished carriers from noncarriers of the detrimental allele, Met66.

For completion we also repeated the analyses coding the variables by the number of minor alleles for all genes except *BDNF*, because there was insufficient number of cases (see Table 1 for the number of participants per number of minor allele).

### Intellectual Enrichment

Education, bilingualism, leisure and professional intellectual activities are some of the activities associated with a protective effect against cognitive decline ^36^. Track-HD recorded main profession, education level using the ISCED scale, and pre-morbid IQ using vocabulary tests. Because different tests were used in each country, pre-morbid IQ was standardized within country (NART- 2 in the UK, ANART in Canada, DART in the Netherlands and Mill Hill in France). Occupational cognitive requirements^37^ were estimated from the main profession recorded for each participant (see supplementary methods). These three measures were then standardized and summed to create a composite score of intellectual enrichment^11^. A higher value in the intellectual enrichment score means higher level of education, a more cognitively demanding profession and higher level of estimated pre-morbid IQ.

### Measures of Disease Pathology

Predicted disease severity at the time of recruitment was measured using the CAG by age product (disease burden score (DBS) = age x (CAG – 35.5)^38^). This is a commonly used model of predicted exposure to disease pathology describing the well-established relationship between age and CAG repeat number of the longer allele. The larger the CAG repeat length, the earlier the predicted age of disease onset^39^.

Pathology at baseline and rate of change was quantified using structural MRI measures of caudate volume and total gray matter (GM) volume, which are robust and well-defined markers of brain atrophy in Huntington’s disease^17–20^. Measures of white matter integrity using diffusion weighted imaging, were only introduced at visit 4 in Track-HD, therefore we only focused on GM volume in our study. Whole brain T1-weighted 3D MPRAGE images were acquired at 3T at all four visits (for details of the imaging protocol see^17^). Caudate volume at baseline and longitudinal change was measured using MIDAS’ semi-automated segmentation and the boundary shift integral respectively^19, 40, 41^. Total GM volume at baseline was measured using SPM12. Longitudinal change was measured using a non-linear fluid registration method in MIDAS which produced whole-brain voxel compression maps measuring change from baseline^42^. Voxel compression maps were then convolved with SPM-derived GM maps to generate change in total GM over time. The measures of caudate volume and total GM volume used in all the analyses were transformed to percent of total intracranial volume (TIV), in order to adjust for differences in brain size. TIV was measured at baseline using MIDAS.

In addition to caudate and total GM volume, we also performed exploratory whole-brain analyses using VBM^43^. The GM probability maps at baseline and voxel-compression maps of change from baseline were normalized to a group template space using DARTEL. Normalized images were then smoothed using an 8mm FWHM gaussian kernel. Full details of the MRI methods used have been published previously^44^.

### Statistical Analyses

Statistical analyses were performed using R version 3.6.3 (http://www.r-project.org/) and the packages lmertest (version 3.1-1) and lme4 (version 1.1-21).

To examine the relationship between cognitive function and brain volume with our predictors of interest, we used linear mixed models. Our predictors of interest were intellectual enrichment and genetic polymorphisms. All models were fitted using maximum likelihood estimation (ML) and correlated random intercept and slope. All models included as covariates age, DBS and study site, and their interaction with visit, as well as sex (main effect only, because the sex by visit interaction term did not improve model fit and was therefore dropped). Models with cognitive function as outcome also included the use of antipsychotic medication as a covariate (see supplementary methods). Time was modelled in years of follow-up as approximated by annual visit number. Based on previously published analyses of the Track-HD data^21^ we included quadratic effects of time in models with cognitive function and total GM volume as the outcome. Age and DBS were mean-centered (Table 1).

Our hypotheses tested whether our variables of interest significantly predicted cognitive function or brain volume at baseline and change over time. We used likelihood ratio tests to assess the covariate-adjusted significance of predictors on the outcome variables. We also tested for the significance of the interaction of genetic predictors with intellectual enrichment in the same way. In all the analyses, we visually inspected model residual distributions to assess plausible normality. No outliers were identified. Significance was established using 2-sided p-values and applying Bonferroni correction to control type I error rate when multiple measures were used to test a hypothesis.

For VBM analyses a binary GM mask was created using the mean normalized images from all Huntington’s disease gene-carriers. This was used in all analyses. Statistical maps were thresholded at two-tailed p < 0.001 uncorrected at voxel-level and p < 0.05 family-wise error (FWE) corrected at cluster-level.

## Data Availability

Track-HD data are available upon request after appropriate data use agreements are signed from the study funder, the CHDI Foundation. Please direct inquiries to.

## Results

### Intellectual enrichment

Previous research showed that participants with early stage Huntington’s disease with higher intellectual enrichment have better cognitive performance than those with lower intellectual enrichment^12^. Furthermore amongst pre-manifest gene-carriers who are closer to predicted disease onset those with high intellectual enrichment have slower rate of cognitive decline than those with low intellectual enrichment^11^.

In our cohort of participants with pre-manifest and early stage Huntington’s disease, intellectual enrichment predicted mean global cognitive function and there was also a significant three-way interaction between intellectual enrichment, DBS and time (i.e. annual visit number) on global cognitive function (both p < 0.001; Table 2 and Supplementary Table 3). In agreement with previous studies, the estimate for the main effect of intellectual enrichment was positive such that participants with high intellectual enrichment had better cognitive function than those with lower) intellectual enrichment. More specifically, for average DBS and age, the mean estimates (95% CI) were -3.14 SD (-3.66, -2.62) and -5.57 (-6.10, -5.04) for high (1SD above mean) and low (1SD below mean) intellectual enrichment respectively. The contrast estimate (SE) for high vs low = 2.43 SD (0.36), t-value(229) = 6.744, p-value < 0.001.

**Table 2:** Association between intellectual enrichment with cognitive function

| Model | Df | AIC | Chisq | P-value |
| --- | --- | --- | --- | --- |
| **Intellectual Enrichment** |  |  |  |  |
| Null model | 20 | 3151.6 |  |  |
| + Intellectual Enrichment | 21 | 3112.3 | 41.3 | <0.001 |
| + Intellectual Enrichment by Visit | 22 | 3113.7 | 0.6 | 0.452 |
| + Intellectual Enrichment by DBS | 23 | 3111.9 | 3.8 | 0.050 |
| + Intellectual Enrichment by DBS by Visit | 24 | 3101.4 | 12.5 | <0.001 |

The null model for intellectual enrichment included age at baseline, site (3 dummy variables) and DBS at baseline with their interaction with Visit, and main effects of sex and use of antipsychotic medication. A quadratic term was also included for Visit, as well as its interaction with DBS.

The estimate for the three-way interaction between DBS, intellectual enrichment and time was in the opposite direction from the main effect; in individuals with low DBS those with high intellectual enrichment declined slower than those with low intellectual enrichment, however in individuals with high DBS, those with high intellectual enrichment declined faster than those with lower intellectual enrichment, despite having better performance at baseline (Figure 1A and Supplementary Figure 1). More specifically, for participants with low DBS (1 SD below mean) those with high intellectual enrichment declined slower than those with lower intellectual enrichment (for 258.9 DBS slope estimate (95% CI) = 0.141 (-0.007, 0.289) and -0.102 (-0.268, 0.065) for high and low intellectual enrichment respectively; contrast estimate (SE) for high vs low intellectual enrichment was 0.243 (0.119), t-value(224) = 2.043, p-value = 0.042). In contrast, an individual with 407.4 DBS (1SD above mean) and 1SD above mean intellectual enrichment had faster cognitive decline than an individual with the same DBS, but 1SD below mean intellectual enrichment (slope estimate (95% CI) = –0.741 (-0.917, -0.565) and -0.413 (- 0.556, -0.270) for high and low intellectual enrichment respectively; contrast estimate (SE) high vs low = -0.327 (0.110), t-value(247) = -2.971, p-value = 0.003). To aid with interpretation of this finding we repeated the same analysis replacing DBS with group as an ordered factor, coding for manifest and pre-manifest individuals. There was a significant main effect of intellectual enrichment, however the group by visit by intellectual enrichment interaction was not significant in this case (Supplementary Table 4). Our results therefore show that there is a strong positive effect of intellectual enrichment on global cognitive function at baseline. It is unclear what is driving the significant 3-way interaction between intellectual enrichment, DBS and visit and whether it is a reliable finding. The lack of a significant 3-way interaction with group suggests that it may not driven by disease stage.

**Figure 1:**
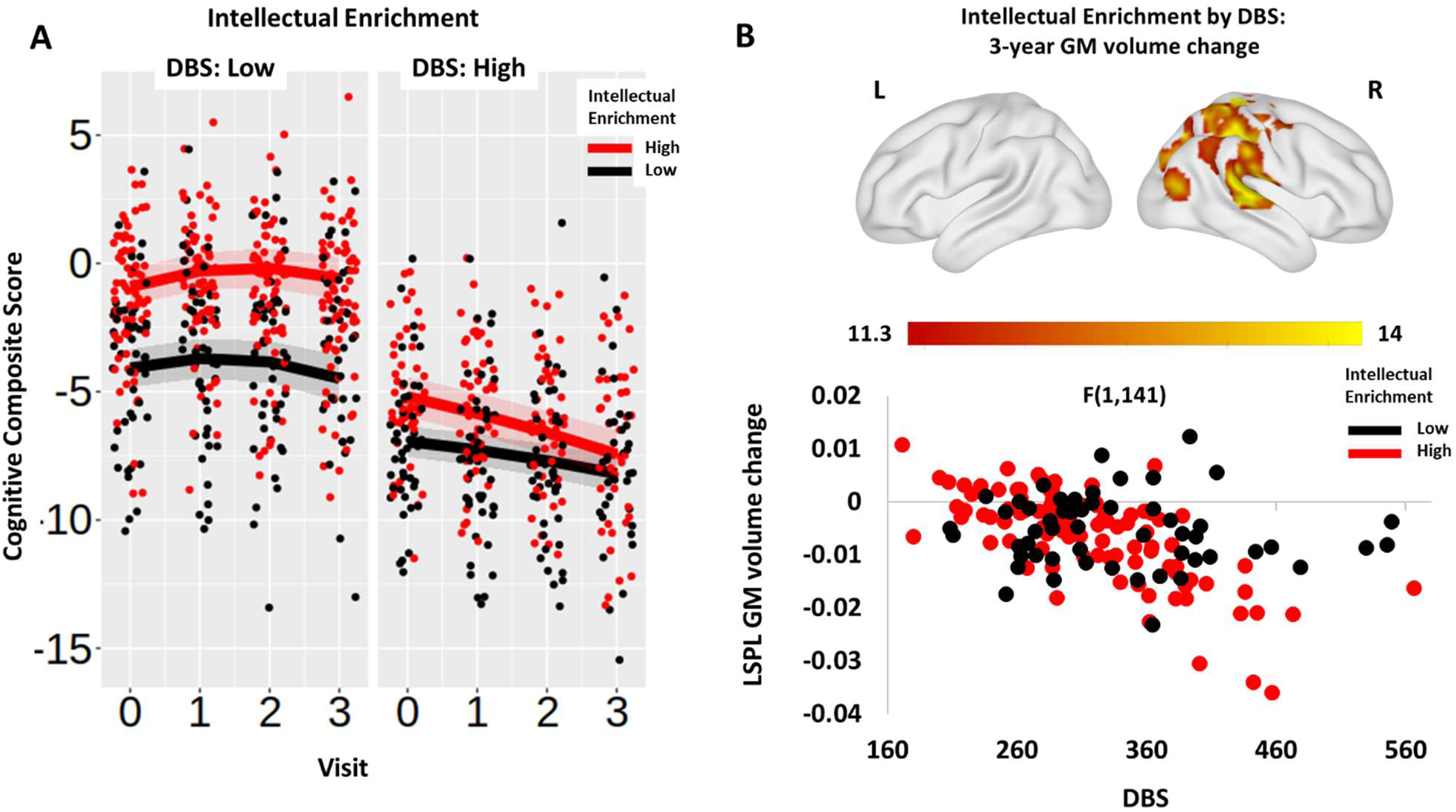
Association between intellectual enrichment and DBS with global cognitive function (A) and 3-year GM volume change (B). In (A) regression lines are generated from the model at high (1SD above mean; red) and low (1SD below mean; black) intellectual enrichment. For visualization purposes results are split into high (above mean) and low (below mean) DBS. Bands around the regression lines are 95% confidence intervals. Datapoints show the raw data residualized against age, site, sex and use of antipsychotic medication, and have been jittered to minimize overlap. In (B) significant clusters are overlaid on the ICBM152 template mesh (top). Maps are thresholded at p < 0.001 uncorrected at voxel level and p < 0.05 family-wise error (FWE) corrected at cluster-level. Shown in a scatter plot (bottom) are the extracted values averaged across the significant cluster. For visualization purposes data are grouped by high (above mean - red) and low (below mean - black) intellectually enrichment. Datapoints show the raw data residualized against age, DBS, site, sex and TIV. LSPL = left superior parietal lobe.

To further understand the mechanism by which intellectual enrichment influences global cognitive function, we next examined its relationship with brain volume, measured by caudate and total GM volume over 3 years. There was a significant interaction between intellectual enrichment and DBS on caudate volume at baseline (p_bon_ = 0.024; Supplementary Table 5 and Supplementary Table 6), such that intellectual enrichment had a positive association with caudate volume in Huntington’s disease gene-carriers far from predicted disease onset (i.e. low DBS), but this effect was attenuated or reversed as the disease progressed (Supplementary Figure 2A). More specifically, a participant with 258.9 DBS (1SD below mean) and high intellectual enrichment had larger caudate volume at baseline compared to a participant with the same DBS, but low intellectual enrichment (mean estimate (95% CI) = 0.0645 (0.0464, 0.0826) and 0.0291 (0.0097, 0.0486) for high and low intellectual enrichment respectively; contrast estimate (SE) high vs low = 0.0354 (0.0139), t-value(215) = 2.534, p-value = 0.012). However, there was no difference in participants with high DBS (for 407.4 DBS contrast estimate (SE) for high vs low intellectual enrichment was -0.0141 (0.135), t-value(215) = -1.044, p-value = 0.298). As previously we repeated the same analysis replacing DBS with group as an ordered factor, coding for manifest vs pre-manifest individuals. There was no significant group by intellectual enrichment interaction on caudate volume at baseline (Supplementary Table 7). It is therefore unclear what is driving the intellectual enrichment by DBS interaction and whether it is reliable. There were no significant main effects or interactions with total GM volume (all p_bon_ > 0.1; Supplementary Table 5 and Supplementary Figure 2B).

Lastly, we performed exploratory, whole-brain analyses using VBM to identify whether there are specific brain regions that showed an effect of intellectual enrichment or an interaction between intellectual enrichment and DBS. There was no significant main effect of intellectual enrichment on volume or volume change anywhere in the brain, but there was a significant interaction with DBS. Participants with low DBS and high intellectual enrichment had larger GM volume at baseline in the right putamen, the thalamus and the right superior temporal gyrus compared to individuals with similar DBS but low intellectual enrichment (Supplementary Table 8 and Supplementary Figure 3). In addition, the rate of GM atrophy over 3 years was faster in individuals with high intellectual enrichment and high DBS in a cluster extending from the right post-central gyrus to the right superior temporal gyrus ventrally and to the superior parietal lobe and the right precuneus caudally (Figure 1B and Supplementary Table 8). As previously we repeated the same analyses replacing DBS with group. There were no brain regions that showed a significant interaction between intellectual enrichment and group for baseline volume or 3 year change.

### Genetic polymorphisms

We next examined the relationship between cognitive function and five genetic polymorphisms linked to genes known to affect cognitive function in Huntington’s disease: *MSH3, FAN1*, *MAPT, BDNF* and *COMT*. We did not find a significant association between *FAN1*, *MAPT, BDNF* and *COMT* variants and global cognitive function at baseline or change over time after Bonferroni correction for five multiple comparisons (all p_bon_ > 0.068; Table 3). In agreement with previous analyses of disease progression in the same cohort ^25^ and recent research^15^, *MSH3* was a significant predictor of global cognitive function at baseline and change (both p_bon_ = 0.045; Table 3 and Supplementary Table 9). More specifically, participants with one or more 3a alleles in *MSH3* had better cognitive function at baseline and slower cognitive decline compared to non-carriers of 3a alleles (for average age and DBS slope (95% CI) = -0.329 (-0.434, -0.223) and -0.118 (-0.241, 0.004) for non-carriers and carriers respectively; contrast estimate (SE) non-carriers vs carriers = -0.21 (0.081), t-value(213) = -2.587, p-value = 0.010; Figure 2A and Supplementary Figure 4). We repeated the analyses coding for the number of alleles in the genes *MSH3*, *FAN1*, *COMT* and *MAPT*. There was no change in the results (see Supplementary Table 10), i.e. only *MSH3* was a significant predictor of cognitive function at baseline and change over time.

**Figure 2:**
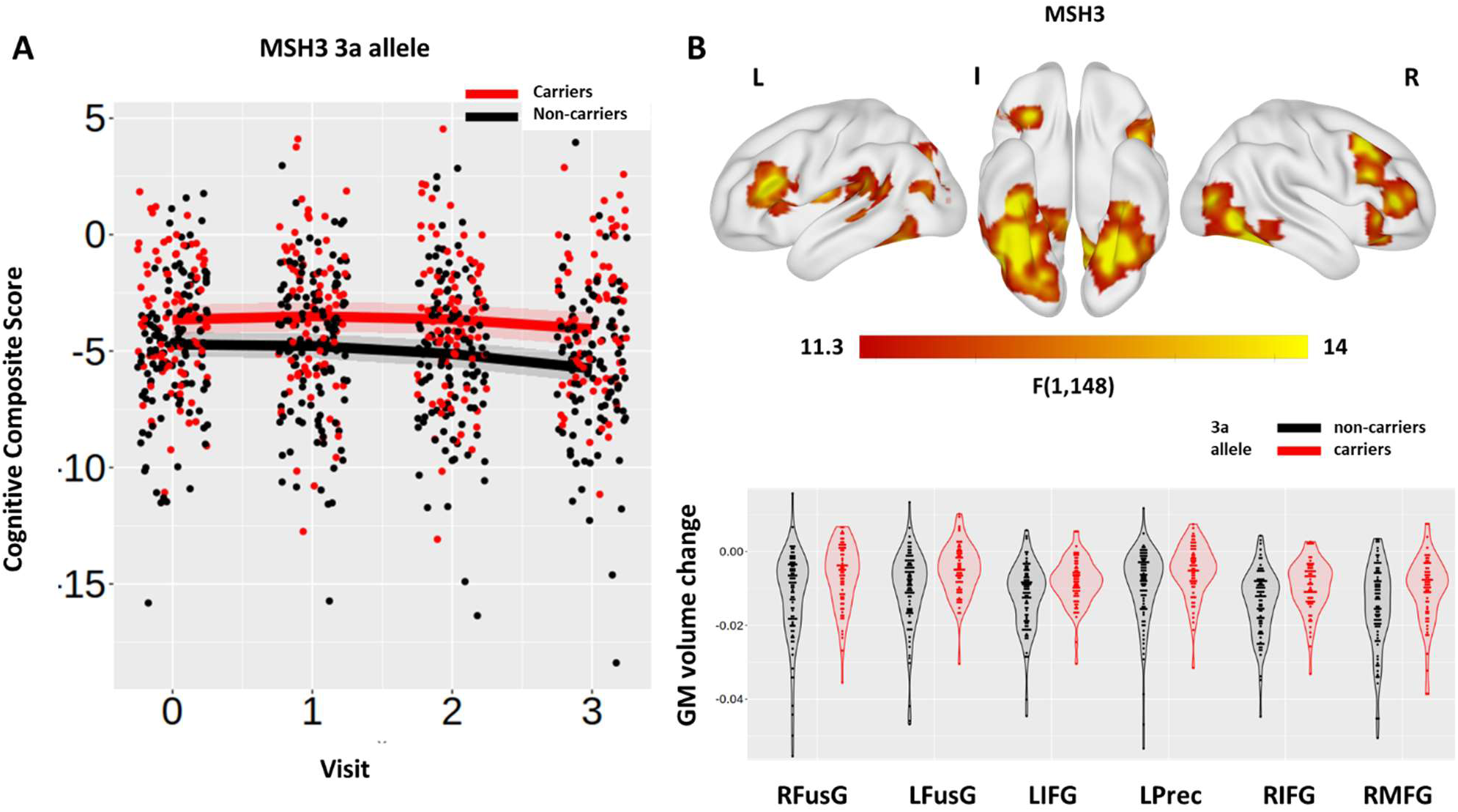
Association between the *MSH3* predictor with global cognitive function (A) and 3-year GM volume change (B). In (A) regression lines show the predicted effect of carrying (red) and not carrying (black) the 3a allele. Bands around the regression lines are 95% confidence intervals. Datapoints show the raw data residualized against age, site, sex and use of antipsychotic medication, and have been jittered to minimize overlap. In (B) significant clusters are overlaid on the ICBM152 template mesh (top). Maps are thresholded at p < 0.001 uncorrected at voxel level and p < 0.05 family-wise error (FWE) corrected at cluster-level. Shown in violin plots (bottom) are the extracted values averaged across the significant clusters for carriers (red) and non-carriers (black) of the 3a allele. Individual datapoints are shown in black dots. Datapoints show the raw data residualized against age, DBS, site, sex and TIV. RFusG and LFusG= right and left fusiform gyrus; LIFG and RIFG= left and right inferior frontal gyrus; LPrec = left precuneus; RMFG = right middle frontal gyrus.

**Table 3:** Association between genetic polymorphisms and cognitive function

| Model | Df | AIC | Chisq | P-value | P-value<br>Bonferroni cor. |
| --- | --- | --- | --- | --- | --- |
| **MSH3** |  |  |  |  |  |
| Null model | 20 | 3139.0 |  |  |  |
| + MSH3 | 21 | 3134.2 | 6.8 | 0.009 | 0.045 |
| + MSH3 by Visit | 22 | 3129.3 | 6.8 | 0.009 | 0.045 |
| + MSH3 by DBS | 23 | 3131.1 | 0.2 | 0.649 | 1 |
| + MSH3 by DBS by Visit | 24 | 3133.1 | 0.0 | 0.850 | 1 |
| **FAN1** |  |  |  |  |  |
| Null model | 20 | 3109.3 |  |  |  |
| + FAN1 | 21 | 3110.7 | 0.6 | 0.4223 | 1 |
| + FAN1 by Visit | 22 | 3112.6 | 0.0 | 0.895 | 1 |
| + FAN1 by DBS | 23 | 3113.1 | 1.6 | 0.213 | 1 |
| + FAN1 by DBS by Visit | 24 | 3113.4 | 1.7 | 0.193 | 0.964 |
| **MAPT** |  |  |  |  |  |
| Null model | 20 | 3109.3 |  |  |  |
| + MAPT | 21 | 3108.8 | 2.6 | 0.110 | 0.550 |
| + MAPT by Visit | 22 | 3110.8 | 0.0 | 0.988 | 1 |
| + MAPT by DBS | 23 | 3107.2 | 5.5 | 0.019 | 0.094 |
| + MAPT by DBS by Visit | 24 | 3109.2 | 0.0 | 0.947 | 1 |
| **COMT** |  |  |  |  |  |
| Null model | 20 | 3109.3 |  |  |  |
| + COMT | 21 | 3110.8 | 0.5 | 0.488 | 1 |
| + COMT by Visit | 22 | 3111.2 | 1.6 | 0.201 | 1 |
| + COMT by DBS | 23 | 3112.6 | 0.5 | 0.462 | 1 |
| + COMT by DBS by Visit | 24 | 3112.2 | 2.4 | 0.120 | 0.601 |
| **BDNF** |  |  |  |  |  |
| Null model | 20 | 3109.3 |  |  |  |
| + BDNF | 21 | 3111.2 | 0.1 | 0.706 | 1 |
| + BDNF by Visit | 22 | 3107.1 | 6.1 | 0.014 | 0.068 |
| + BDNF by DBS | 23 | 3109.0 | 0 | 0.889 | 1 |
| + BDNF by DBS by Visit | 24 | 3110.9 | 0.1 | 0.741 | 1 |

The null model included age at baseline, site (3 dummy variables), DBS at baseline with their interaction with Visit, sex and use of antipsychotic medication. A quadratic term was also included for Visit, as well as its interaction with DBS. P-values were corrected for 5 independent comparisons using Bonferroni correction.

**Table 4:** Interaction between intellectual enrichment and genetic polymorphisms on cognitive function

| Model | Df | AIC | Chisq | P-value | P-value Bonferroni cor. |
| --- | --- | --- | --- | --- | --- |
| **MSH3** |  |  |  |  |  |
| Null model | 24 | 2931.8 |  |  |  |
| + MSH3 by Intellectual Enrichment | 25 | 2933.1 | 0.7 | 0.404 | 1 |
| + MSH3 by Intellectual Enrichment by Visit | 26 | 2933.9 | 1.1 | 0.286 | 1 |
| **FAN1** |  |  |  |  |  |
| Null model | 24 | 2910.6 |  |  |  |
| + FAN1 by Intellectual Enrichment | 25 | 2912.4 | 0.2 | 0.679 | 1 |
| + FAN1 by Intellectual Enrichment by Visit | 26 | 2914.4 | 0.0 | 0.833 | 1 |
| **MAPT** |  |  |  |  |  |
| Null model | 24 | 2911.5 |  |  |  |
| + MAPT by Intellectual Enrichment | 25 | 2913.5 | 0.0 | 0.895 | 1 |
| + MAPT by Intellectual Enrichment by Visit | 26 | 2915.5 | 0.0 | 0.894 | 1 |
| **COMT** |  |  |  |  |  |
| Null model | 24 | 2910.2 |  |  |  |
| + COMT by Intellectual Enrichment | 25 | 2912.2 | 0.0 | 0.960 | 1 |
| + COMT by Intellectual Enrichment by Visit | 26 | 2914.0 | 0.2 | 0.630 | 1 |
| **BDNF** |  |  |  |  |  |
| Null model | 24 | 2907.6 |  |  |  |
| + BDNF by Intellectual Enrichment | 25 | 2902.1 | 7.5 | 0.006 | 0.031 |
| + BDNF by Intellectual Enrichment by Visit | 26 | 2903.8 | 0.2 | 0.627 | 1 |

The null model included intellectual enrichment, the genetic polymorphism, age at baseline, site (3 dummy variables), DBS at baseline with their interaction with Visit, sex and use of antipsychotic medication. A quadratic term was also included for Visit, as well as its interaction with DBS. P-values were corrected for 5 independent comparisons using Bonferroni correction.

We next examined the relationship between *MSH3* polymorphisms and brain volume to further understand the mechanism by which *MSH3* may influence global cognitive function. *MSH3* had a significant effect on total GM volume rate of change (i.e. *MSH3* by Visit interaction; p_bon_ = 0.001; Supplementary Table 5, Supplementary Table 11 and Supplementary Figure 5).

Huntington’s disease gene carriers with one or more 3a alleles in *MSH3* had slower rate of total GM atrophy over 3 years compared to non-carriers (for average age and DBS slope (95% CI) = -0.181 (-0.202, -0.160) and -0.122 (-0.148, -0.097) for non-carriers and carriers respectively; contrast estimate (SE) for non-carriers vs carriers = -0.0586 (0.0168), t-value(196) = -3.486, p < 0.001).

Exploratory VBM analyses examined the main effect of *MSH3* on GM volume across the whole-brain. There was a significant effect of *MSH3* on GM volume at baseline in the right middle temporal gyrus, the right inferior occipital gyrus and the left post-central gyrus (Supplementary Table 8 and Supplementary Figure 6). There was also a significant effect of *MSH3* on GM volume change in the fusiform gyrus bilaterally, the inferior frontal gyrus bilaterally, the right middle frontal gyrus and the left precuneus (Supplementary Table 8 and Figure 2B). Lastly there was an interaction between *MSH3* and DBS on GM volume change in the left inferior frontal gyrus, the left supplementary motor area and the right superior temporal gyrus (Supplementary Table 8 and Supplementary Figure 7). The effect of *MSH3* was positive in all cases such that 3a allele carriers (across all participants or for those with high DBS) had higher volume and slower rate of GM atrophy compared to non-carriers. Our results therefore suggest that carrying the 3a allele in *MSH3* supported preserved cognitive function and was associated with a slower rate of neurodegeneration.

### Gene-environment interaction

Previous research in ageing and dementia has shown that environmental factors, including intellectual enrichment, interact with genetic polymorphisms in order to predict cognitive function^7, 8^. To test this hypothesis in our study we examined the interaction between all five genetic polymorphisms and intellectual enrichment on cognitive function and decline. *BDNF* was the only gene which significantly interacted with intellectual enrichment to predict global cognitive function at baseline (p_bon_ = 0.031 corrected for five tests; Table 3 and Supplementary Table 12). There was a positive interaction between *BDNF* and intellectual enrichment on global cognitive function at baseline, such that the effect of intellectual enrichment on cognitive function was stronger for Met66 allele carriers than non-carriers. More specifically, the estimates for Met66 allele carriers with low (1SD below mean) and high intellectual enrichment (1SD above mean) were: estimate (95% CI) = -6.31 (-7.20, -5.42) and -2.57 (-3.41, -1.73) respectively; contrast estimate (SE) for high vs low intellectual enrichment = 3.74 (0.598), t-value(216) = 6.249, p-value < 0.001. Similarly, for Met66 allele non-carriers, with low (1SD below mean) and high intellectual enrichment (1SD above mean) estimate (95% CI) = -5.08 (-5.75, -4.42) and -3.40 (-4.10, -2.71) respectively; contrast estimate (SE) for high vs low intellectual enrichment = 1.68 (0.483), t-value(214) = 3.486, p-value < 0.001. The difference between high and low intellectual enrichment was greater for Met66 allele carriers than non-carriers suggesting that the Met66 allele moderates the effect of intellectual enrichment on cognitive function (contrast estimate (SE) = -2.06 (0.768), t-value(215) = -2.678, p-value = 0.008; Figure 3).

**Figure 3:**
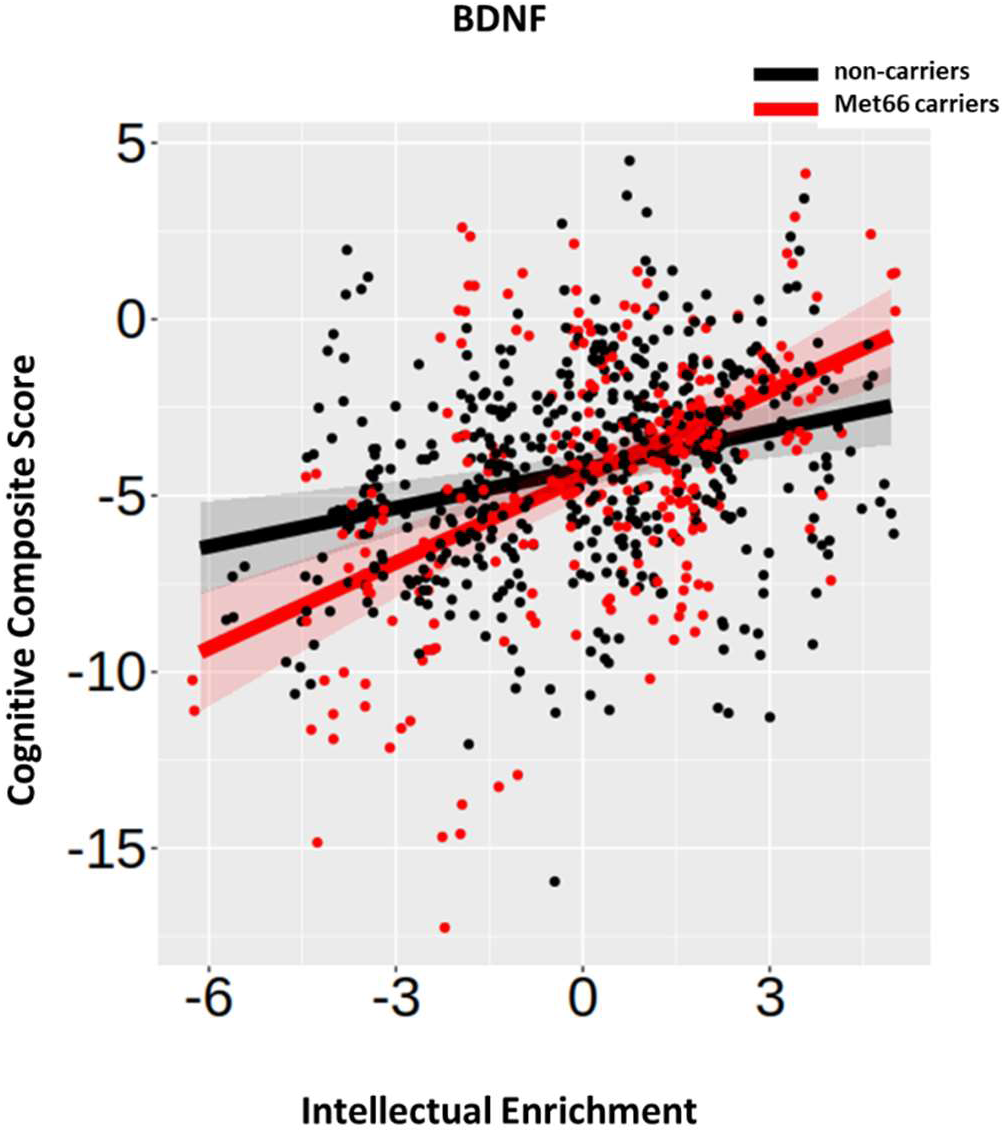
Association between the *BDNF* predictor and intellectual enrichment with global cognitive function. Regression lines show the predicted effect of carrying (red) and not carrying (black) the Met66 allele. Bands around the regression lines are 95% confidence intervals. Datapoints show the raw data residualized against age, site, sex and use of antipsychotic medication.

To understand the mechanism by which *BDNF* interacts with intellectual enrichment to impact cognitive function, we examined the interaction between *BDNF* and intellectual enrichment on brain volume. The effect of the interaction of *BDNF* with intellectual enrichment was significant, but small, on both caudate and total GM volume rate of change (both p_bon_ < 0.03 corrected for two tests; Supplementary Table 13-15). The difference in rate of volume change between individuals with high and low intellectual enrichment was positive in Met66 allele carriers, but negative for non-carriers. In more detail, for Met66 allele carriers with high (1SD above mean) and low intellectual enrichment (1SD below mean) estimate (95% CI) = -0.0094 (-0.0105, - 0.0082) and -0.0110 (-0.0123, -0.0097) respectively; contrast estimate (SE) = 0.0017 (0.0009), t- value(228) =1.900, p-value = 0.059. For Met66 allele non-carriers with high (1SD above mean) and low intellectual enrichment (1SD below mean) estimate (95% CI) = -0.0103 (-0.0112, - 0.0094) and -0.0094 (-0.0102, -0.0085) respectively; contrast estimate (SE) = -0.0010 (0.0007), t-value(205) = -1.480, p-value = 0.140. The difference between high and low intellectual enrichment was positive for Met66 allele carriers, but negative for non-carriers, suggesting that the Met66 allele moderates the effect of intellectual enrichment on caudate atrophy rate (contrast estimate (SE) = -0.0026 (0.0011), t-value(221) = -2.406, p-value = 0.017; Supplementary Figure 8).

This was similar for total GM volume. For Met66 allele carriers with high (1SD above mean) and low intellectual enrichment (1SD below mean) estimate (95% CI) = -0.129 (-0.167, -0.090) and -0.206 (-0.248, -0.1635) respectively; contrast estimate (SE) = 0.0769 (0.0289), t-value(214) = 2.658, p-value = 0.008. For Met66 allele non-carriers with high (1SD above mean) and low intellectual enrichment (1SD below mean) estimate (95% CI) = -0.179 (-0.209, -0.1484) and - 0.132 (-0.161, -0.1020) respectively; contrast estimate (SE) = -0.0469 (0.0218), t-value(192) =- 2.154, p-value = 0.0324. The difference between high and low intellectual enrichment was positive for Met66 allele carriers, but negative for non-carriers, suggesting that the Met66 allele moderates the effect of intellectual enrichment on total GM atrophy rate (contrast estimate (SE) = -0.124 (0.0362), t-value(207) = -3.421, p-value = 0.008; Supplementary Figure 9).

Exploratory whole-brain VBM analyses examined the interaction between *BDNF* and intellectual enrichment on GM volume at baseline and change over time, but we did not identify any significant clusters.

## Discussion

The aim of our study was to examine the role of intellectual enrichment and genetic polymorphisms on cognitive function and brain structure in Huntington’s disease. Our results highlight the complexity of the interplay between environmental and genetic factors on behaviour and brain structure. Intellectual enrichment and genetic variation in *MSH3* are independently associated with global cognitive function and brain structure, whereas intellectual enrichment interacts with *BDNF* to attenuate the deleterious effect of the Met66 polymorphism.

In more detail, we replicated previous findings showing that intellectual enrichment was associated with better global cognitive function ^11^. In the present work, we further show that in participants with high DBS (in our study mean DBS was 333.2) those with high intellectual enrichment had faster rate of decline over 3 years (0.36 SD annualized change in global cognitive function), than those with lower intellectual enrichment (Figure 1A). The faster rate of decline in participants with high DBS narrowed the difference in baseline cognitive performance between individuals with low and high intellectual enrichment. Similarly, we found that high intellectual enrichment predicted accelerated atrophy in posterior cortical regions of the right hemisphere in participants with high DBS. Therefore, it appears that as DBS increases the protective effect of intellectual enrichment on cognition decreases. However, when replacing DBS with group the 3-way interaction between intellectual enrichment, group and visit was not significant in relation to cognitive function or brain volume. Given the very strong association between DBS and group, it is therefore unclear what is driving the significant interaction between intellectual enrichment and DBS and whether it is a reliable finding.

Further insights regarding the mechanism by which intellectual enrichment affects cognitive function are provided by the interaction between intellectual enrichment and *BDNF* gene variation. The difference in cognitive function between individuals with low and high intellectual enrichment was greater in Met66 allele carriers, than non-carriers. Comparing the blood expression levels of *BDNF* between 22 controls and 62 manifest gene carriers from this cohort using RNAseq, we previously found the HD gene-carriers had lower levels of *BDNF* expression than controls (p- value = 0.04266^45^). Post-mortem studies have also identified reduced BDNF levels in the striatum of Huntington’s disease patients. However, it is unclear whether this is due to defects in the delivery of cortical BDNF^46^ or to the response in the striatum^31^. It is possible that carrying the Met66 allele exacerbates existing defects in the BDNF pathway. Previous research in animal models of Huntington’s disease showed that such defects can be rescued by environmental enrichment^47^. Our results are in broad agreement with these findings and suggest that intellectual enrichment potentially counteracts the detrimental effect of the Met66 allele in both cognitive function and brain structure (striatum and total GM). Our findings are also in agreement with previous studies in ageing showing a significant interaction between intellectual enrichment and *BDNF* to predict cognitive function and decline in healthy older adults^7, 32^.

Intellectual enrichment did not interact with any of the other genetic polymorphisms we examined, whereas the only genetic predictor with a significant effect on cognitive function was variation in *MSH3*. A recent study^15^ has identified *MSH3* as a modifier of cognitive function in HD, while we have previously shown in this cohort that variation in *MSH3*, specifically carrying a 3a allele, has a protective effect on a composite score of disease progression which included cognitive and psychomotor function^22, 25^. It is currently hypothesized that *MSH3* is introducing an expansion of the *HTT* CAG repeat in the process of repair. Greater expansion is associated with earlier disease onset and faster progression^48, 49^, whereas carrying the 3a allele is associated with reduced expression of *MSH3* and therefore reduced somatic expansion and slower progression^25^. In agreement with this finding, in the present work we further show that carrying the 3a allele in *MSH3* was associated with slower GM atrophy across different regions in the cortex, including the inferior temporal and inferior frontal gyri. The absence of a significant effect in the striatum is notable given previous work which showed that there is large somatic expansion in both the cortex (temporal, occipital and prefrontal cortex) and the striatum^50^. This finding could be explained by the fact that all analyses were adjusted for differences in DBS and suggest that carrying the 3a allele does not explain additional variance in striatal atrophy. Lastly, it is important to note that the protective effect of carrying the 3a allele in *MSH3* is a result of reducing the expression of *MSH3*, and there is no evidence that it supports neuroprotective mechanisms. The effect of MSH3 on cognitive function and brain volume cannot therefore be interpreted as brain maintenance.

Similar to *MSH3,* variation in *FAN1* (rs3512) has also been implicated in somatic expansion instability^49^ and has been previously shown to predict delayed age of onset in Huntington’s disease^24, 34^. *FAN1* overexpression reduces CAG repeat expansion in human cell modes^51^, however in our study there was no significant effect of rs2140734 (or rs3512) on cognitive function. The reason for the contradictory findings is unclear. It could be due to lack of statistical power given the relatively small sample size, however recent work suggests a differential effect of *FAN1* on motor rather than cognitive function^15^.

Lastly, in contrast to previous studies in Huntington’s disease, we did not find any evidence for an association between the genes *COMT* and *MAPT* and cognitive function. A previous study^14^ showed that *COMT* Val158 allele carriers had slower cognitive decline compared to Met158 homozygotes in manifest Huntington’s disease patients. The number of Met158 homozygotes was low in our cohort (43 out of 229; 19%) and our cohort included both pre-manifest and patients at early stages of the disease, which could explain the contradictory findings. Variation in the *MAPT* gene has also been previously shown to predict cognitive decline in Huntington’s disease^16^, such that H1 homozygotes had slower decline in cognitive function compared to H2 carriers. However, this effect was opposite to what has been previously reported for Parkinson’s disease^52^, and had a small effect size (r = -0.14). In our study we did not find strong evidence for an association between *MAPT* and cognitive function, it is therefore currently unclear whether *MAPT* plays a role in cognitive function in Huntington’s disease.

In summary, we have shown that cognitive function in Huntington’s disease is affected by an interplay between genetic and environmental factors. Cognitive decline is slower in carriers of the 3a allele in the gene *MSH3,* supported by the slowing of GM volume atrophy in the cortex, in agreement with the role of *MSH3* in somatic expansion instability. Intellectual enrichment also appears to have a protective effect on cognitive function at baseline independent of disease stage. Importantly, we also observed a significant interaction between intellectual enrichment and the *BDNF* gene polymorphism, whereby intellectual enrichment counteracted the detrimental effect of carrying the Met66 allele on cognitive function and brain structure, in agreement with the role of intellectual enrichment in enhancing brain trophic support. Future research is now needed to develop and evaluate intellectual enrichment interventions in Huntington’s disease and measure its impact in both behaviour and brain structure.

## Data Availability

Track-HD data are available upon request after appropriate data use agreements are signed from
the study funder, the CHDI Foundation. Please direct inquiries to.

## Acknowledgements

TRACK-HD investigators Peter Kraus, Rainer Hoffman, Alan Tobin, Beth Borowsky, S. Keenan, Kathryn B. Whitlock, Sarah Queller, Colin Campbell, Chiachi Wang, Eric Axelson, Hans Johnson, Tanka Acharya, Dave M. Cash, Chris Frost, Rebecca Jones, Caroline Jurgens, Ellen P. ‘t Hart, Jeroen van der Grond, Marie-Noelle N. Witjes-Ane, Raymund A.C. Roos, Eve M. Dumas, Simon J.A. van den Bogaard, Cheryl Stopford, David Craufurd, Jenny Callaghan, Natalie Arran, Diana D. Rosas, S. Lee, W Monaco, Alison O’Regan, Cassie Milchman, Ellen Frajman, Izelle Labuschagne, Julie Stout, Melissa Campbell, Sophie C. Andrews, Natalie Bechtel, Ralf Reilmann, Stefan Bohlen, Chris Kennard, Claire Berna, Stephen Hicks, Alexandra Durr, Cristophe Pourchot, Eric Bardinet, Kevin Nigaud, Romain Valabrègue, Stephane Lehericy, Cecilia Marelli, Celine Jauffret, Damian Justo, Blair Leavitt, Joji Decolongon, Aaron Sturrock, Alison Coleman, Rachelle Dar Santos, Aakta Patel, Claire Gibbard, Daisy Whitehead, Ed Wild, Gail Owen, Helen Crawford, Ian Malone, Nayana Lahiri, Nick C. Fox, Nicola Z. Hobbs, Roger Ordidge, Tracey Pepple, Joy Read, Miranda J. Say, Bernhard Landwehrmeyer

## Abbreviations

BDNF: brain-derived neurotrophic factor
CFI: comparative fit index
CI: confidence interval
COMT: catechol-O-methyl transferase
CSF: cerebrospinal fluid
DART: Dutch adult reading test
DARTEL: diffeomorphic anatomical registration through exponentiated lie algebra
DBS: disease burden score
DF: degrees of freedom
FAN1: fancd2- and fanci-associated nuclease 1
FWE: family-wise error
FWHM: full-width at half maximum
GM: gray matter
HADS: hospital anxiety and depression scale
HD: Huntington’s disease
HTT: Huntingtin
IQ: intelligence quotient
ISCED: international standard classification of education
MAPT: microtubule-associated protein tau
MIDAS: medical image display and analysis software
ML: maximum likelihood
MPRAGE: magnetization-prepared rapid acquisition with gradient echo
MSH3: MutS homolog 3
(A)NART: (American) national adult reading test
RMSEA: root mean square error of approximation
SD: standard deviation
SDMT: symbol digit modalities test
SE: standard error
SEM: structural equation modelling
SNP: single nucleotide polymorphism
SPM: statistical parametric mapping
SRMR: standardised root mean residual
TIV: total intracranial volume
TREM2: triggering receptor expressed on myeloid cells 2
UHDRS TMS: unified Huntington’s disease rating scale total motor score
VBM: voxel-based morphometry

## STROBE statement: Reporting guidelines checklist for cohort, case-control and cross-sectional studies

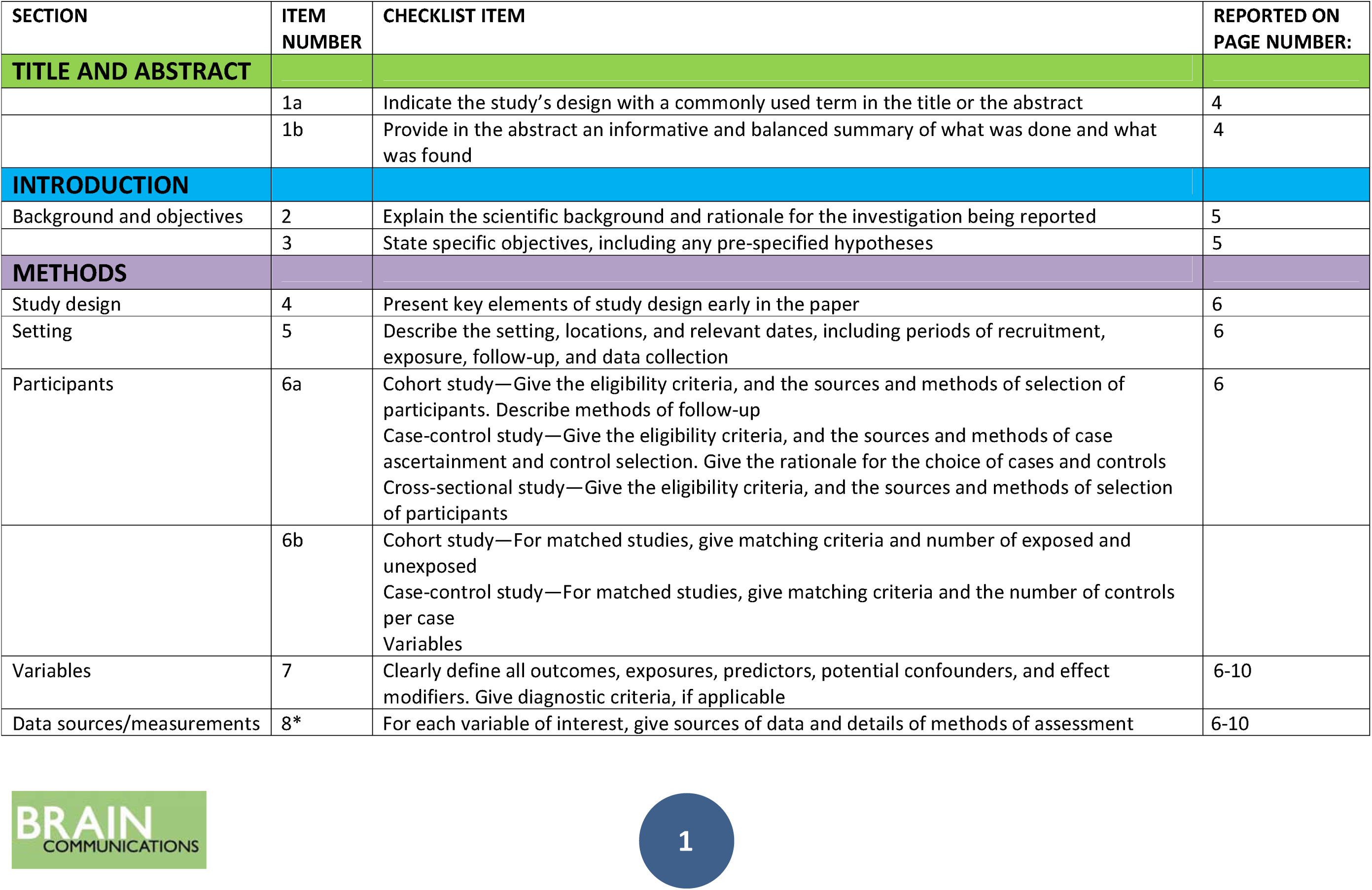
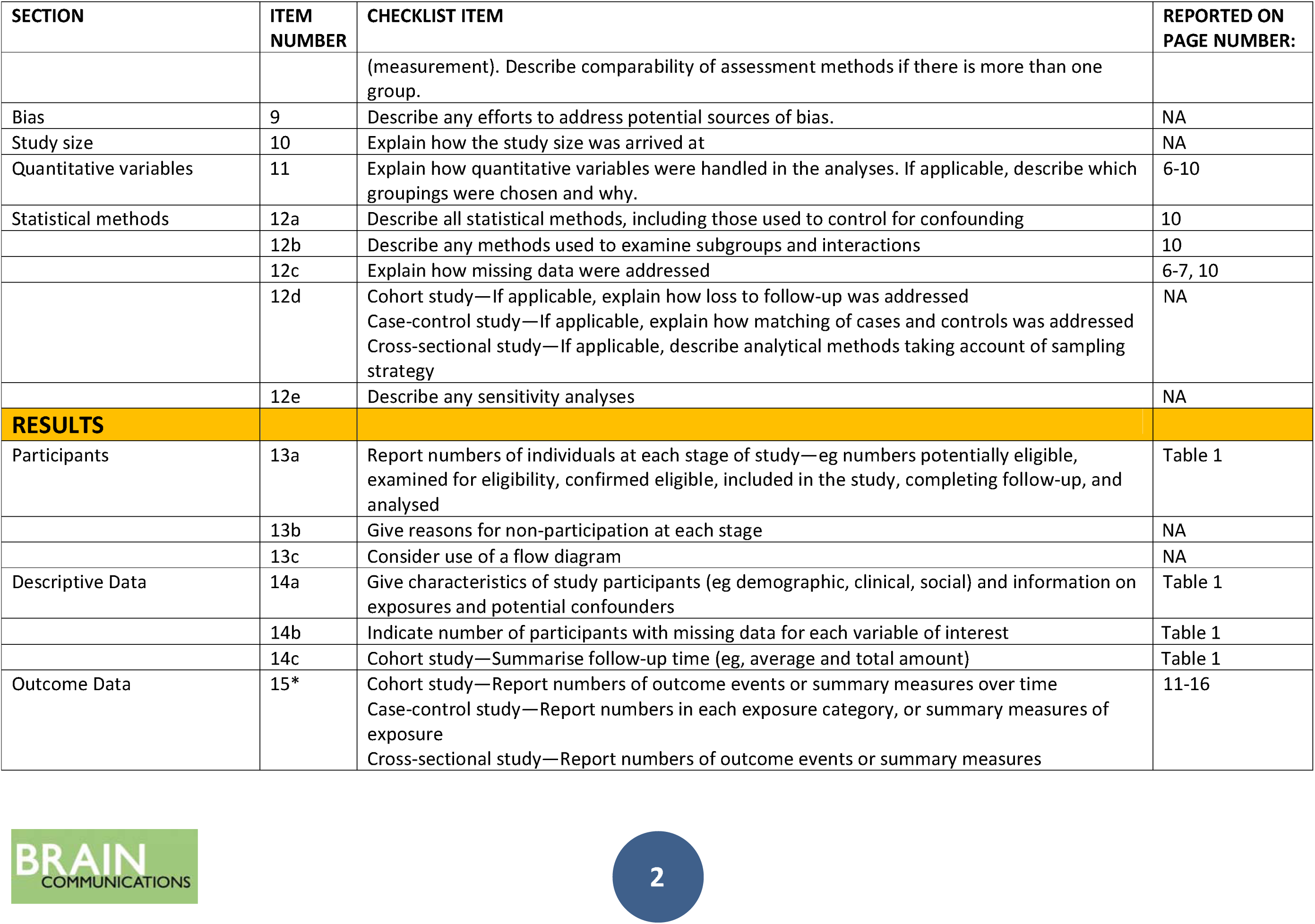
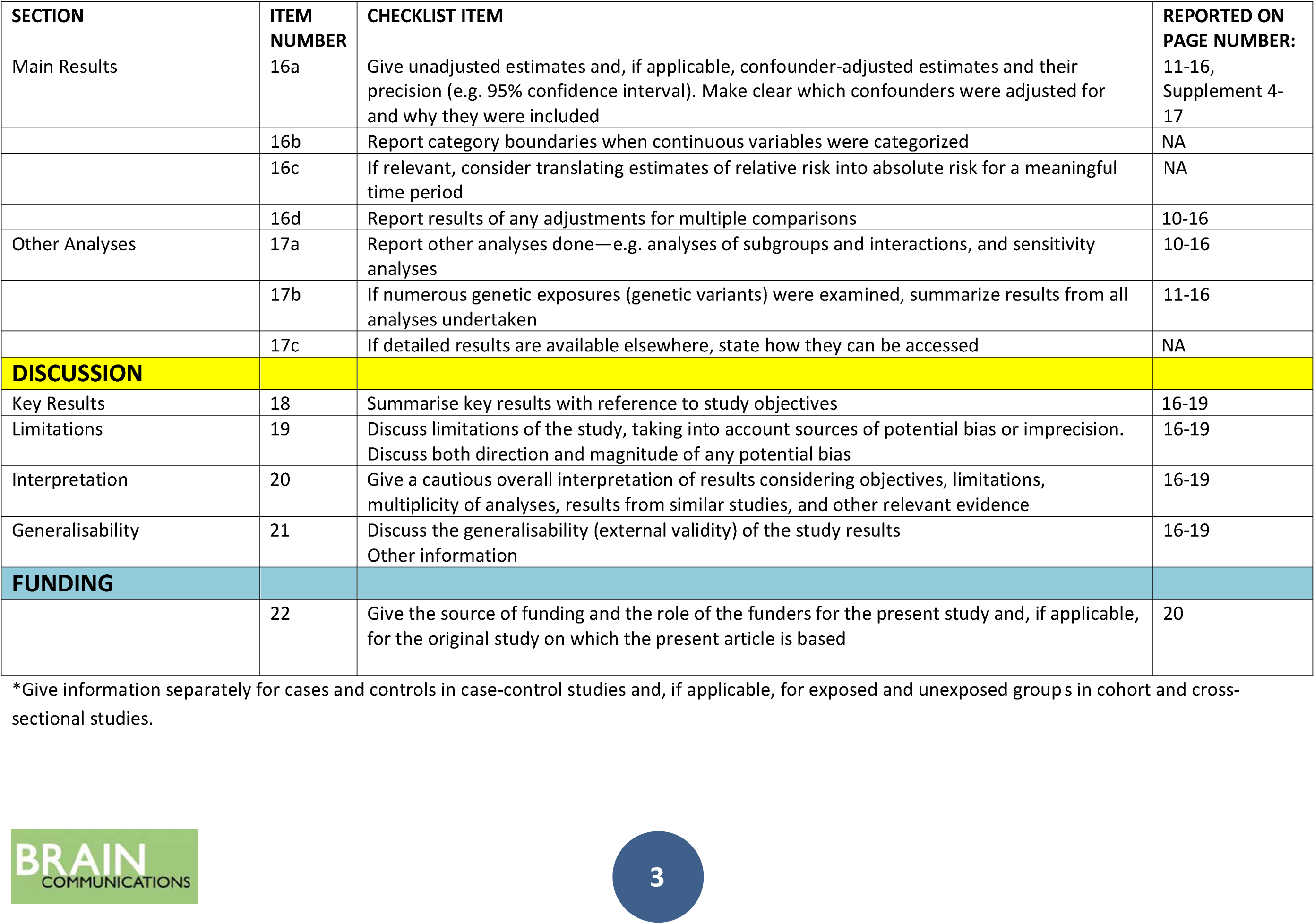

## Supplementary Methods

### Occupational cognitive requirements

As a measure of occupational intellectual activity we used the occupational cognitive requirements of an individual’s main occupation. We used the approach developed by Pool and colleagues^1^ using O*NET scores, which was shown to predict cognitive function in healthy older adults. Primary occupation at the Visit of assessment or main occupation, if an individual had retired, was matched to O*NET-SOC occupation titles (version 23.2^2^). We then created a measure of occupational requirements by averaging the scores from the 10 cognitive related O*NET items: processing information, thinking creatively, judging the qualities of things, services or people, evaluating information to determine compliance with standards, analyzing data or information, making decisions and solving problems, updating and using relevant knowledge, developing objectives and strategies, scheduling work and activities, organizing, planning and prioritizing. Higher scores indicate higher cognitive requirements. Non-paid occupations such as housewife or students are not included in O*NET and were not given a score. They were 3 such cases in our cohort. In 7 cases it was not possible to find a clear match with O*NET occupation titles, because the occupation recorded was unclear, e.g. financial expert. These cases were therefore excluded for the analyses.

### Control risk factors

The use of antipsychotic mediation has been associated with cognitive impairment^3^. In our cohort, there were 34 (14.8%) participants on antipsychotic medication on their baseline visit. The antipsychotic medication they were prescribed were: olanzapine, risperidone, aripiprazole, quetiapine, sulpiride, tiapride, amisulpride, tetrabenazine, paliperidone, clozapine and pimozide. The use of antipsychotic medication had a negative effect on cognitive function in our cohort (p < 0.001; parameter estimate (95% CI) = -0.855 (-1.30, - 0.41); supplementary table 15). We therefore included it as a control variable in all the models with cognitive function as an outcome measure.

The presence of depression has also been associated with impaired cognitive function^4^. There were 28 (12.6%) participants who scored greater than 7 on the HADS^5^, indicating the presence of depression, whereas for 6 participants there was no data on the HADS. The presence of depression was not a significant predictor of cognitive function in our cohort (p > 0.05; see supplementary table 15).

Lastly, we examined ApoE to test for concurrent risk of Alzheimer’s dementia (AD) in e4 allele carriers in our cohort^6^. To determine APoE alleles in the Track-HD cohort rs429358 and rs7412 were imputed in Minimac4 using HRC.r1-1.GRCh37. A binary variable was created contrasting carriers and non-carriers of the detrimental e4 allele. There were 5 ambiguous cases in our cohort, which were either e2/e4 or e1/e3, and 16 participants with missing data. These were excluded from our analyses. In total, there were 60 (28.8%) e4 allele carriers in our cohort. Analyses testing for the effect of carrying the e4 allele on cognitive function in our cohort showed that it was not a significant predictor (p > 0.09; supplementary Table 15) and was therefore not included in any analyses.

## Supplementary Tables

**Supplementary Table 1:**
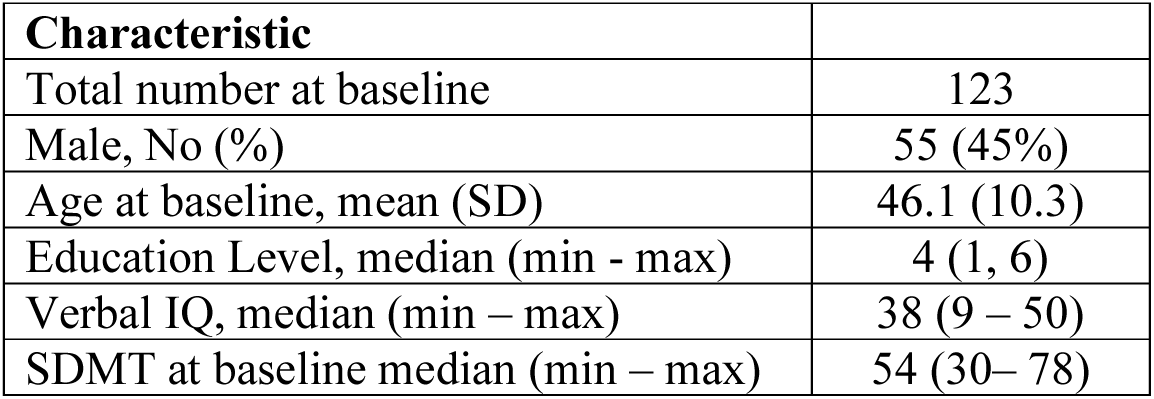
Control group demographics

**Supplementary Table 2:**
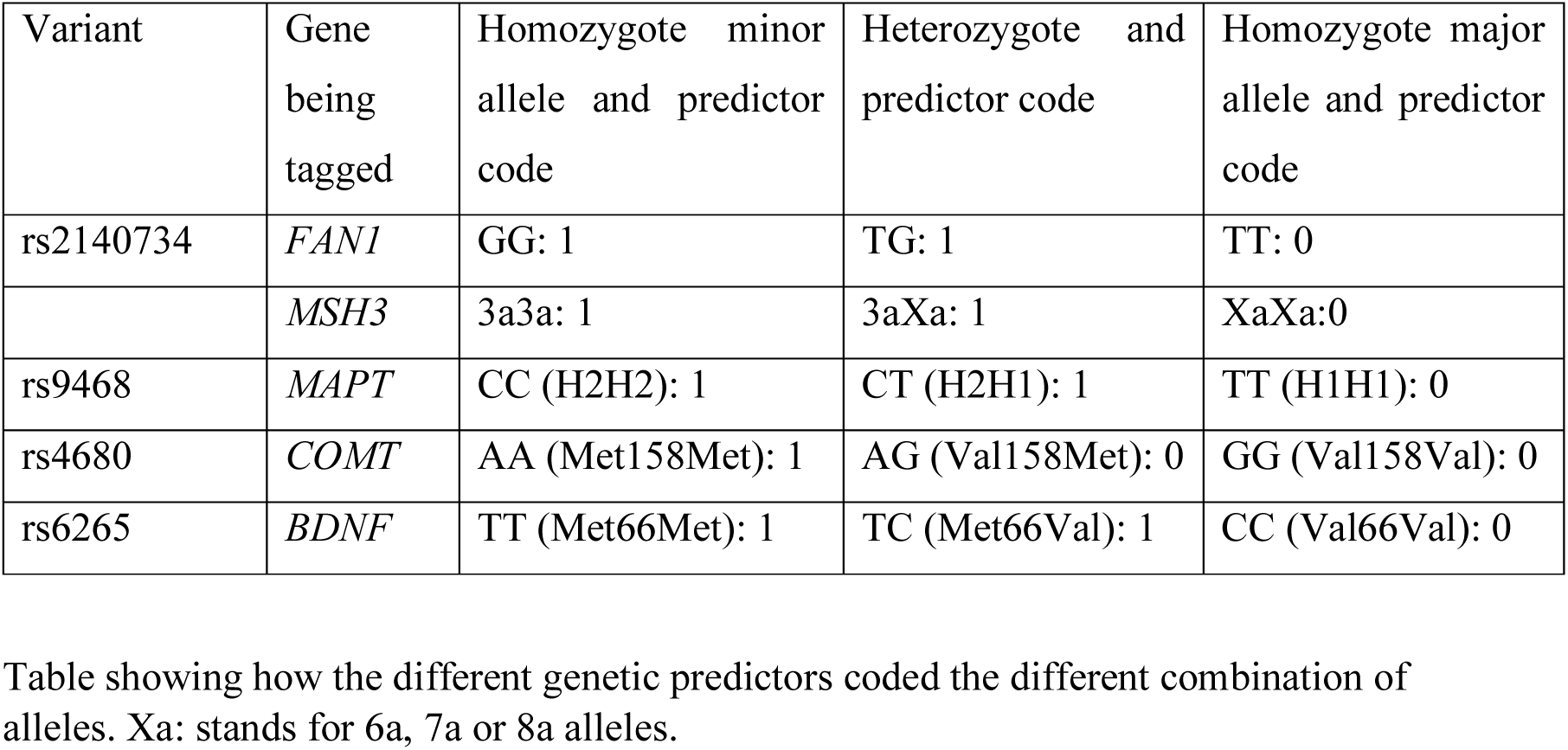
Coding of binary genetic predictors

**Supplementary Table 3:** Regression coefficients for intellectual enrichment and DBS model with cognitive function as outcome variable

| **Parameter** | **Estimate** | **SE** | **DF** | **T-value** | **P-value** |
| --- | --- | --- | --- | --- | --- |
| Intercept | -4.19400 | 0.37410 | 220.9 | -11.212 | <0.001 |
| Visit | 0.23080 | 0.12320 | 604.9 | 1.873 | 0.061 |
| Visit<sup>2</sup> | -0.14450 | 0.03317 | 425.0 | -4.357 | <0.001 |
| DBS | -0.02394 | 0.00246 | 232.8 | -9.746 | <0.001 |
| Age | -0.13680 | 0.01735 | 221.4 | -7.889 | <0.001 |
| Site - London | 1.33400 | 0.48640 | 217.2 | 2.742 | 0.007 |
| Site - Paris | 0.19150 | 0.49300 | 221.7 | 0.388 | 0.698 |
| Site - Vancouver | 0.59340 | 0.48070 | 217.6 | 1.234 | 0.218 |
| Sex - Male | -0.58060 | 0.35320 | 217.4 | -1.644 | 0.102 |
| Antipsychotic medication | -0.84600 | 0.21840 | 742.8 | -3.874 | <0.001 |
| Intellectual Enrichment | 0.52510 | 0.07620 | 217.7 | 6.890 | <0.001 |
| Intellectual Enrichment by DBS | -0.00207 | 0.00101 | 217.9 | -2.052 | 0.041 |
| Visit by Intellectual Enrichment | -0.01167 | 0.01713 | 213.4 | -0.681 | 0.497 |
| Visit by Intellectual Enrichment by DBS | -0.00083 | 0.00023 | 225.1 | -3.596 | <0.001 |
| Visit by Age | -0.00588 | 0.00376 | 201.8 | -1.562 | 0.120 |
| Visit by DBS | -0.00802 | 0.00143 | 525.8 | -5.626 | <0.001 |
| Visit by Site - London | -0.19400 | 0.10720 | 205.7 | -1.810 | 0.072 |
| Visit by Site – Paris | -0.01218 | 0.10760 | 203.7 | -0.113 | 0.910 |
| Visit by Site - Vancouver | -0.21800 | 0.10620 | 212.0 | -2.053 | 0.041 |
| Visit<sup>2</sup> by DBS | 0.00141 | 0.00045 | 429.9 | 3.119 | 0.002 |

**Supplementary Table 4:** Regression coefficients for model with intellectual enrichment and group terms, with cognitive function as outcome variable

| **Parameter** | **Estimate** | **SE** | **DF** | **T-value** | **P-value** |
| --- | --- | --- | --- | --- | --- |
| Intercept | -3.947837 | 0.353470 | 222.5 | -11.169 | <0.001 |
| Visit | 0.307414 | 0.125318 | 602.5 | 2.453 | <0.001 |
| Visit<sup>2</sup> | -0.152926 | 0.033323 | 427.9 | -4.589 | <0.001 |
| Group | -3.009590 | 0.269153 | 234.1 | -11.182 | <0.001 |
| Age | -0.052558 | 0.018763 | 219.1 | -2.801 | 0.006 |
| Site - London | 1.466529 | 0.466580 | 218.4 | 3.143 | 0.002 |
| Site - Paris | 0.430566 | 0.474181 | 222.7 | 0.908 | 0.365 |
| Site - Vancouver | 0.380672 | 0.459623 | 218.7 | 0.828 | 0.408 |
| Sex - Male | -0.820579 | 0.338105 | 218.8 | -2.427 | 0.016 |
| Antipsychotic medication | -0.865641 | 0.218431 | 747.2 | -4.589 | <0.001 |
| Intellectual Enrichment | 0.507534 | 0.074440 | 218.7 | 6.818 | <0.001 |
| Intellectual Enrichment by Group | 0.010694 | 0.103148 | 218.4 | 0.104 | 0.918 |
| Visit by Intellectual Enrichment | -0.009345 | 0.018207 | 217.7 | -0.513 | 0.608 |
| Visit by Intellectual Enrichment by Group | -0.004113 | 0.025552 | 216.4 | -0.161 | 0.872 |
| Visit by Age | 0.003780 | 0.004437 | 205.9 | 0.852 | 0.395 |
| Visit by Group | -0.580852 | 0.152329 | 550.1 | -3.813 | <0.001 |
| Visit by Site - London | -0.201426 | 0.112600 | 212.0 | -1.789 | 0.075 |
| Visit by Site – Paris | 0.001570 | 0.113417 | 210.6 | 0.014 | 0.989 |
| Visit by Site - Vancouver | -0.269283 | 0.110897 | 217.8 | -2.428 | 0.016 |
| Visit<sup>2</sup> by Group | 0.067339 | 0.047191 | 428.4 | 1.427 | 0.154 |

**Supplementary Table 5:**
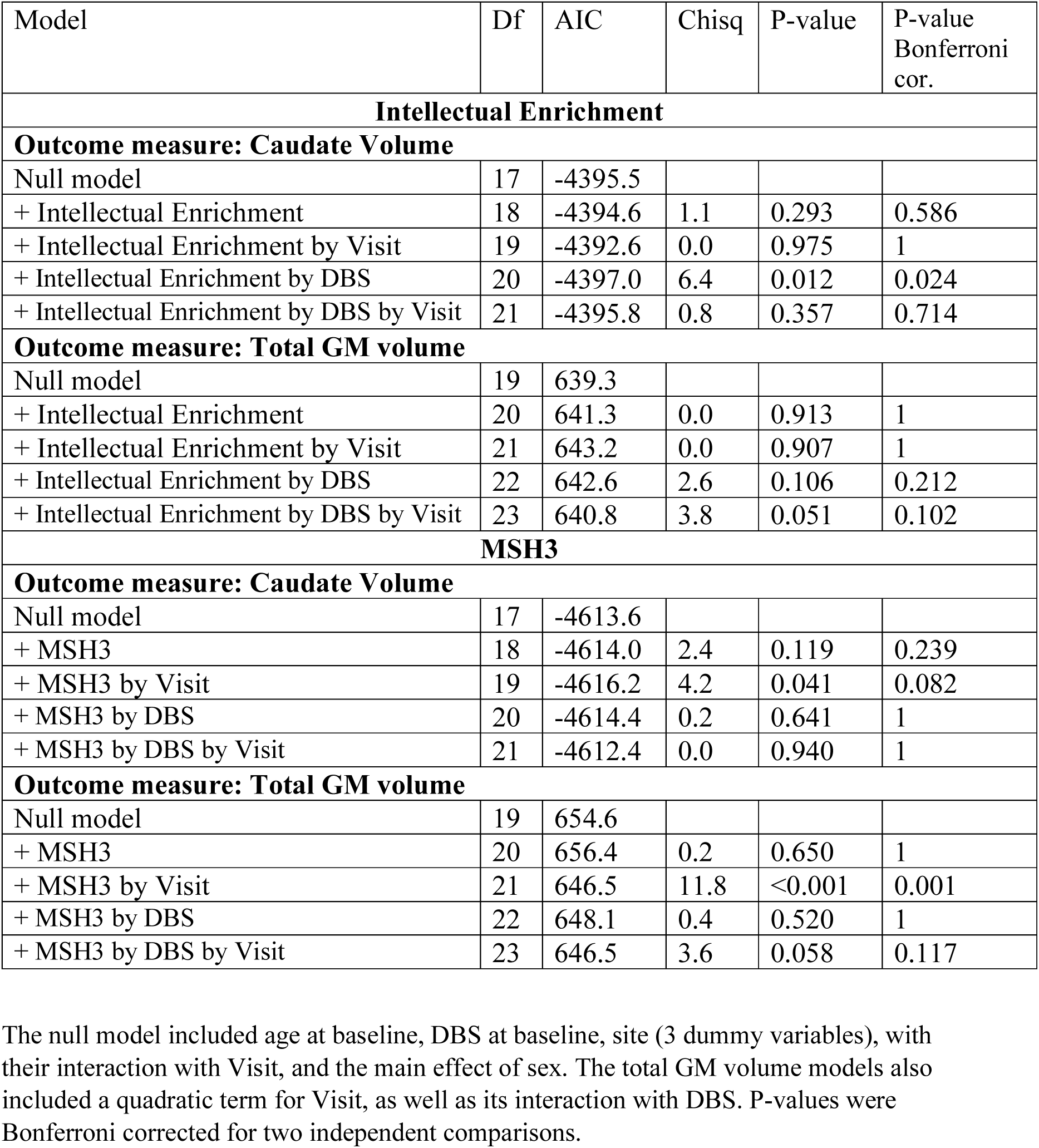
Association between intellectual enrichment and *MSH3* with brain volume

| Model | Df | AIC | Chisq | P-value | P-value Bonferroni cor. |
| --- | --- | --- | --- | --- | --- |
| **Intellectual Enrichment** |  |  |  |  |  |
| **Outcome measure: Caudate Volume** |  |  |  |  |  |
| Null model | 17 | -4395.5 |  |  |  |
| + Intellectual Enrichment | 18 | -4394.6 | 1.1 | 0.293 | 0.586 |
| + Intellectual Enrichment by Visit | 19 | -4392.6 | 0.0 | 0.975 | 1 |
| + Intellectual Enrichment by DBS | 20 | -4397.0 | 6.4 | 0.012 | 0.024 |
| + Intellectual Enrichment by DBS by Visit | 21 | -4395.8 | 0.8 | 0.357 | 0.714 |
| **Outcome measure: Total GM volume** |  |  |  |  |  |
| Null model | 19 | 639.3 |  |  |  |
| + Intellectual Enrichment | 20 | 641.3 | 0.0 | 0.913 | 1 |
| + Intellectual Enrichment by Visit | 21 | 643.2 | 0.0 | 0.907 | 1 |
| + Intellectual Enrichment by DBS | 22 | 642.6 | 2.6 | 0.106 | 0.212 |
| + Intellectual Enrichment by DBS by Visit | 23 | 640.8 | 3.8 | 0.051 | 0.102 |
| **MSH3** |  |  |  |  |  |
| **Outcome measure: Caudate Volume** |  |  |  |  |  |
| Null model | 17 | -4613.6 |  |  |  |
| + MSH3 | 18 | -4614.0 | 2.4 | 0.119 | 0.239 |
| + MSH3 by Visit | 19 | -4616.2 | 4.2 | 0.041 | 0.082 |
| + MSH3 by DBS | 20 | -4614.4 | 0.2 | 0.641 | 1 |
| + MSH3 by DBS by Visit | 21 | -4612.4 | 0.0 | 0.940 | 1 |
| **Outcome measure: Total GM volume** |  |  |  |  |  |
| Null model | 19 | 654.6 |  |  |  |
| + MSH3 | 20 | 656.4 | 0.2 | 0.650 | 1 |
| + MSH3 by Visit | 21 | 646.5 | 11.8 | <0.001 | 0.001 |
| + MSH3 by DBS | 22 | 648.1 | 0.4 | 0.520 | 1 |
| + MSH3 by DBS by Visit | 23 | 646.5 | 3.6 | 0.058 | 0.117 |

The null model included age at baseline, DBS at baseline, site (3 dummy variables), with their interaction with Visit, and the main effect of sex. The total GM volume models also included a quadratic term for Visit, as well as its interaction with DBS. P-values were Bonferroni corrected for two independent comparisons.

**Supplementary Table 6:**
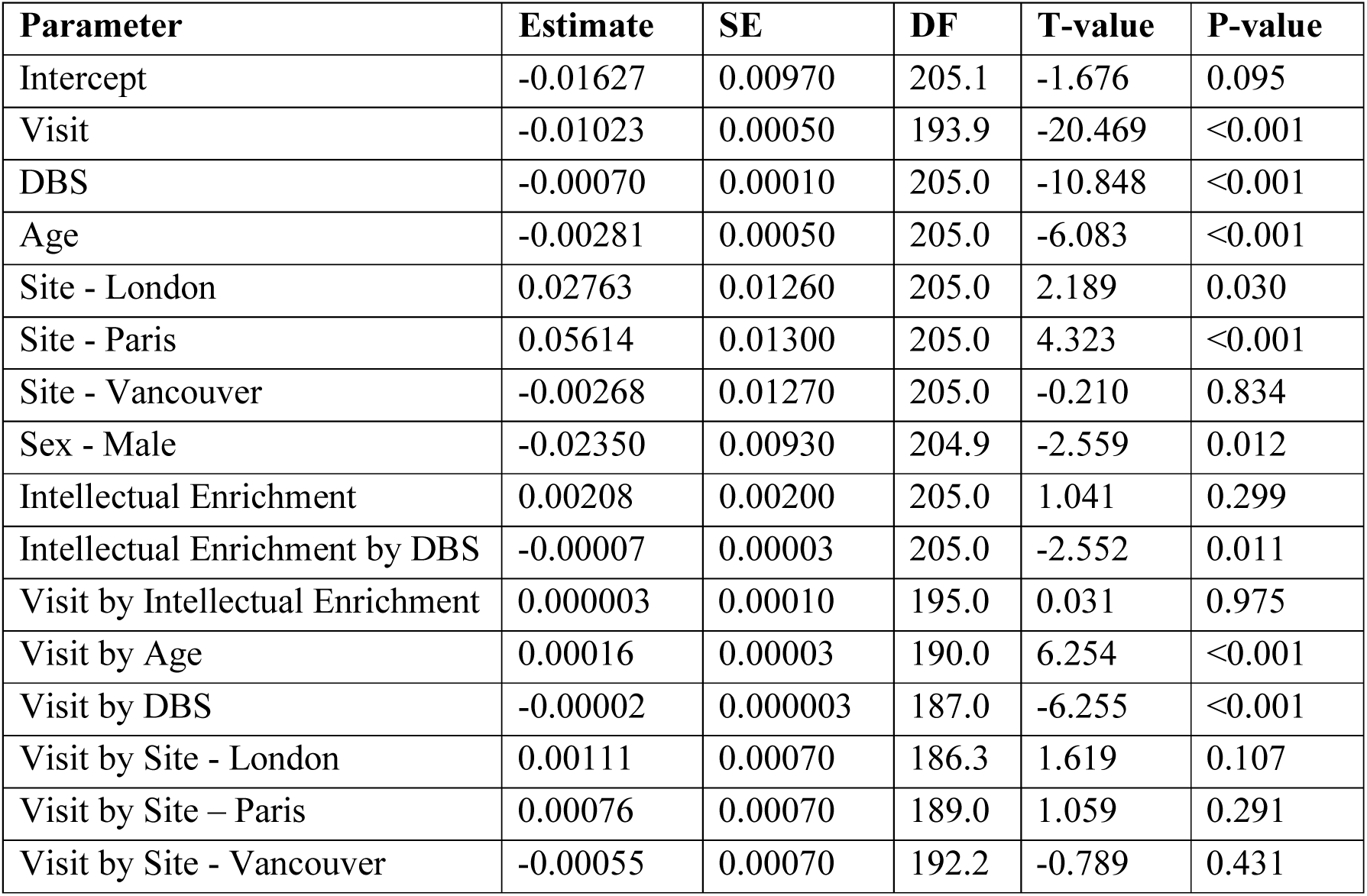
Regression coefficients for intellectual enrichment and DBS model with caudate volume as outcome variable

| **Parameter** | **Estimate** | **SE** | **DF** | **T-value** | **P-value** |
| --- | --- | --- | --- | --- | --- |
| Intercept | -0.01627 | 0.00970 | 205.1 | -1.676 | 0.095 |
| Visit | -0.01023 | 0.00050 | 193.9 | -20.469 | <0.001 |
| DBS | -0.00070 | 0.00010 | 205.0 | -10.848 | <0.001 |
| Age | -0.00281 | 0.00050 | 205.0 | -6.083 | <0.001 |
| Site - London | 0.02763 | 0.01260 | 205.0 | 2.189 | 0.030 |
| Site - Paris | 0.05614 | 0.01300 | 205.0 | 4.323 | <0.001 |
| Site - Vancouver | -0.00268 | 0.01270 | 205.0 | -0.210 | 0.834 |
| Sex - Male | -0.02350 | 0.00930 | 204.9 | -2.559 | 0.012 |
| Intellectual Enrichment | 0.00208 | 0.00200 | 205.0 | 1.041 | 0.299 |
| Intellectual Enrichment by DBS | -0.00007 | 0.00003 | 205.0 | -2.552 | 0.011 |
| Visit by Intellectual Enrichment | 0.000003 | 0.00010 | 195.0 | 0.031 | 0.975 |
| Visit by Age | 0.00016 | 0.00003 | 190.0 | 6.254 | <0.001 |
| Visit by DBS | -0.00002 | 0.000003 | 187.0 | -6.255 | <0.001 |
| Visit by Site - London | 0.00111 | 0.00070 | 186.3 | 1.619 | 0.107 |
| Visit by Site – Paris | 0.00076 | 0.00070 | 189.0 | 1.059 | 0.291 |
| Visit by Site - Vancouver | -0.00055 | 0.00070 | 192.2 | -0.789 | 0.431 |

**Supplementary Table 7:** Regression coefficients for intellectual enrichment and group model with caudate volume as outcome variable

| **Parameter** | **Estimate** | **SE** | **DF** | **T-value** | **P-value** |
| --- | --- | --- | --- | --- | --- |
| Intercept | 0.04489 | 0.01089 | 205.6 | 4.123 | <0.001 |
| Visit | -0.00860 | 0.00056 | 188.0 | -15.329 | <0.001 |
| Group | -0.10450 | 0.01057 | 205.0 | -9.884 | <0.001 |
| Age | -0.00072 | 0.00054 | 205.1 | -1.346 | 0.180 |
| Site - London | 0.03067 | 0.01304 | 205.0 | 2.353 | 0.020 |
| Site - Paris | 0.06290 | 0.01343 | 205.0 | 4.683 | <0.001 |
| Site - Vancouver | -0.01060 | 0.01309 | 205.1 | -0.810 | 0.419 |
| Sex - Male | -0.03177 | 0.00410 | 204.8 | -3.357 | <0.001 |
| Intellectual Enrichment | 0.00288 | 0.00314 | 206.5 | 0.916 | 0.360 |
| Intellectual Enrichment by Group | -0.00160 | 0.00410 | 205.1 | -0.389 | 0.698 |
| Visit by Intellectual Enrichment | 0.00008 | 0.00011 | 194.3 | 0.066 | 0.947 |
| Visit by Age | 0.00021 | 0.00003 | 187.4 | 7.337 | <0.001 |
| Visit by Group | -0.00296 | 0.00057 | 184.4 | -5.192 | <0.001 |
| Visit by Site - London | 0.00121 | 0.00071 | 186.3 | 1.703 | 0.090 |
| Visit by Site – Paris | 0.00091 | 0.00073 | 188.4 | 1.237 | 0.217 |
| Visit by Site - Vancouver | -0.00088 | 0.00072 | 191.3 | -1.231 | 0.220 |

**Supplementary Table 8:**
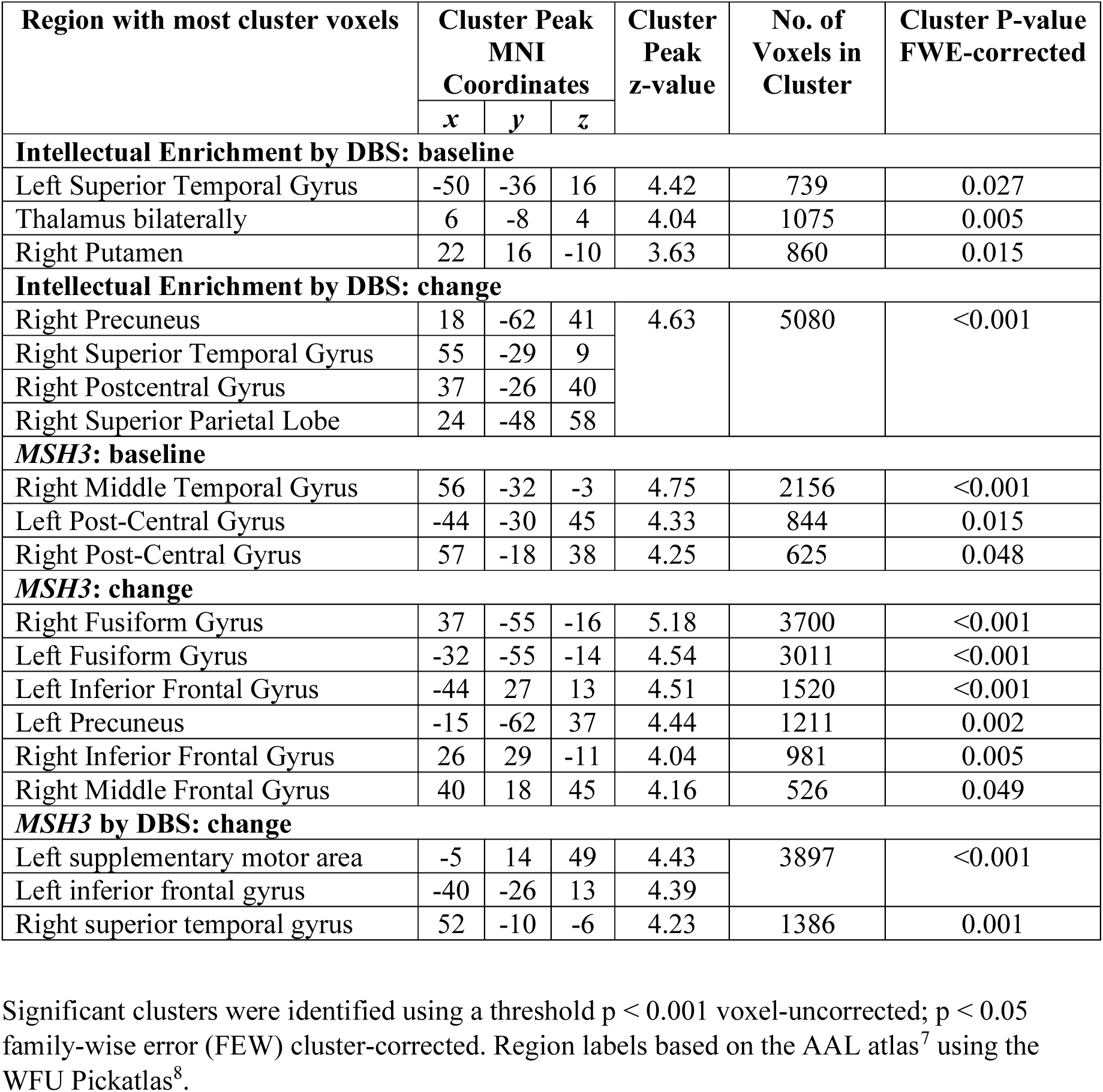
VBM results for intellectual enrichment and *MSH3*

| Region with most cluster voxels | Cluster Peak MNI Coordinates |  |  | Cluster Peak z-value | No. of Voxels in Cluster | Cluster P-value FWE-corrected |
| --- | --- | --- | --- | --- | --- | --- |
|  | *x* | *y* | *z* |  |  |  |
| Intellectual Enrichment by DBS: baseline |  |  |  |  |  |  |
| Left Superior Temporal Gyrus | -50 | -36 | 16 | 4.42 | 739 | 0.027 |
| Thalamus bilaterally | 6 | -8 | 4 | 4.04 | 1075 | 0.005 |
| Right Putamen | 22 | 16 | -10 | 3.63 | 860 | 0.015 |
| Intellectual Enrichment by DBS: change |  |  |  |  |  |  |
| Right Precuneus | 18 | -62 | 41 | 4.63 | 5080 | <0.001 |
| Right Superior Temporal Gyrus | 55 | -29 | 9 |  |  |  |
| Right Postcentral Gyrus | 37 | -26 | 40 |  |  |  |
| Right Superior Parietal Lobe | 24 | -48 | 58 |  |  |  |
| MSH3: baseline |  |  |  |  |  |  |
| Right Middle Temporal Gyrus | 56 | -32 | -3 | 4.75 | 2156 | <0.001 |
| Left Post-Central Gyrus | -44 | -30 | 45 | 4.33 | 844 | 0.015 |
| Right Post-Central Gyrus | 57 | -18 | 38 | 4.25 | 625 | 0.048 |
| MSH3: change |  |  |  |  |  |  |
| Right Fusiform Gyrus | 37 | -55 | -16 | 5.18 | 3700 | <0.001 |
| Left Fusiform Gyrus | -32 | -55 | -14 | 4.54 | 3011 | <0.001 |
| Left Inferior Frontal Gyrus | -44 | 27 | 13 | 4.51 | 1520 | <0.001 |
| Left Precuneus | -15 | -62 | 37 | 4.44 | 1211 | 0.002 |
| Right Inferior Frontal Gyrus | 26 | 29 | -11 | 4.04 | 981 | 0.005 |
| Right Middle Frontal Gyrus | 40 | 18 | 45 | 4.16 | 526 | 0.049 |
| MSH3 by DBS: change |  |  |  |  |  |  |
| Left supplementary motor area | -5 | 14 | 49 | 4.43 | 3897 | <0.001 |
| Left inferior frontal gyrus | -40 | -26 | 13 | 4.39 |  |  |
| Right superior temporal gyrus | 52 | -10 | -6 | 4.23 | 1386 | 0.001 |

Significant clusters were identified using a threshold $p < 0.001$ voxel-uncorrected; $p < 0.05$ family-wise error (FEW) cluster-corrected. Region labels based on the AAL atlas<sup>7</sup> using the WFU Pickatlas<sup>8</sup>.

**Supplementary Table 9:** Regression coefficients for *MSH3* model with cognitive function as outcome variable

| Parameter | Estimate | SE | DF | T-value | P-value |
| --- | --- | --- | --- | --- | --- |
| Intercept | -4.80200 | 0.04485 | 217.3 | -10.707 | <0.001 |
| Visit | 0.21170 | 0.13120 | 576.5 | 1.613 | 0.107 |
| Visit<sup>2</sup> | -0.14060 | 0.03371 | 419.8 | -4.169 | <0.001 |
| DBS | -0.02637 | 0.00276 | 229.1 | -9.546 | <0.001 |
| Age | -0.12280 | 0.01939 | 216.2 | -6.333 | <0.001 |
| Site - London | 1.49100 | 0.55050 | 213.6 | 2.708 | 0.007 |
| Site - Paris | 0.70900 | 0.55300 | 217.4 | 1.282 | 0.201 |
| Site - Vancouver | 0.75470 | 0.55430 | 213.8 | 1.362 | 0.175 |
| Sex - Male | -0.23600 | 0.39550 | 214.0 | -0.597 | 0.551 |
| Antipsychotic medication | -1.07000 | 0.24260 | 714.5 | -4.412 | <0.001 |
| *MSH3* - 3a allele carriers | 1.06500 | 0.39950 | 215.7 | 2.666 | 0.008 |
| Visit by *MSH3* | -0.21030 | 0.07973 | 198.7 | 2.637 | 0.009 |
| Visit by Age | -0.00725 | 0.00389 | 200.6 | -1.865 | 0.064 |
| Visit by DBS | -0.00747 | 0.00147 | 516.9 | -5.095 | <0.001 |
| Visit by Site - London | -0.23530 | 0.11170 | 202.4 | -2.107 | 0.036 |
| Visit by Site – Paris | -0.07850 | 0.11060 | 199.9 | -0.710 | 0.479 |
| Visit by Site - Vancouver | -0.22820 | 0.11260 | 208.2 | -2.028 | 0.044 |
| Visit<sup>2</sup> by DBS | 0.00144 | 0.00046 | 418.9 | 3.106 | 0.002 |

**Supplementary Table 10:** Regression coefficients for *MSH3, FAN1, MAPT* and *COMT* models coding for the number of alleles with cognitive function as outcome variable

| Model | Df | AIC | Chisq | P-value | P-value<br>Bonferroni cor. |
| --- | --- | --- | --- | --- | --- |
| **MSH3** |  |  |  |  |  |
| Null model | 20 | 3139.0 |  |  |  |
| + MSH3 | 21 | 3135.3 | 5.7 | 0.017 | 0.084 |
| + MSH3 by Visit | 22 | 3130.6 | 6.7 | 0.010 | 0.049 |
| + MSH3 by DBS | 23 | 3131.7 | 0.9 | 0.337 | 1 |
| + MSH3 by DBS by Visit | 24 | 3133.7 | 0.0 | 0.855 | 1 |
| **FAN1** |  |  |  |  |  |
| Null model | 20 | 3109.3 |  |  |  |
| + FAN1 | 21 | 3110.4 | 0.9 | 0.936 | 1 |
| + FAN1 by Visit | 22 | 3112.2 | 0.2 | 0.203 | 1 |
| + FAN1 by DBS | 23 | 3113.6 | 0.6 | 0.580 | 1 |
| + FAN1 by DBS by Visit | 24 | 3110.7 | 4.9 | 0.027 | 0.135 |
| **MAPT** |  |  |  |  |  |
| Null model | 20 | 3109.3 |  |  |  |
| + MAPT | 21 | 3108.8 | 2.5 | 0.114 | 0.568 |
| + MAPT by Visit | 22 | 3110.8 | 0.0 | 0.926 | 1 |
| + MAPT by DBS | 23 | 3109.1 | 3.7 | 0.054 | 0.269 |
| + MAPT by DBS by Visit | 24 | 3111.1 | 0.0 | 0.881 | 1 |
| **COMT** |  |  |  |  |  |
| Null model | 20 | 3109.3 |  |  |  |
| + COMT | 21 | 3111.2 | 0.1 | 0.789 | 1 |
| + COMT by Visit | 22 | 3112.7 | 0.5 | 0.461 | 1 |
| + COMT by DBS | 23 | 3113.6 | 1.1 | 0.299 | 1 |
| + COMT by DBS by Visit | 24 | 3115.4 | 0.2 | 0.691 | 1 |

**Supplementary Table 11:** Regression coefficients for *MSH3* model with total GM volume as outcome variable

| **Parameter** | **Estimate** | **SE** | **DF** | **T-value** | **P-value** |
| --- | --- | --- | --- | --- | --- |
| Intercept | 0.99730 | 0.44610 | 222.5 | 2.236 | 0.026 |
| Visit | -0.07065 | 0.01983 | 360.1 | -3.562 | <0.001 |
| Visit<sup>2</sup> | -0.03044 | 0.00385 | 368.1 | -7.914 | <0.001 |
| DBS | -0.01376 | 0.00280 | 206.0 | -4.907 | <0.001 |
| Age | -0.20420 | 0.02022 | 205.0 | -10.100 | <0.001 |
| Site - London | -0.75250 | 0.56470 | 204.6 | -1.333 | 0.184 |
| Site - Paris | -0.65440 | 0.56400 | 205.2 | -1.160 | 0.247 |
| Site - Vancouver | -0.11330 | 0.58070 | 206.7 | -0.195 | 0.846 |
| Sex - Male | -1.58400 | 0.35710 | 206.6 | -4.436 | <0.001 |
| *MSH3* - 3a allele carriers | 0.17100 | 0.36380 | 209.1 | 0.470 | 0.639 |
| Visit by Age | -0.00438 | 0.00085 | 197.8 | -5.135 | <0.001 |
| Visit by DBS | -0.00049 | 0.00019 | 535.3 | -2.565 | 0.011 |
| Visit by Site - London | 0.00604 | 0.02316 | 194.7 | 0.261 | 0.795 |
| Visit by Site – Paris | -0.01008 | 0.02335 | 199.3 | -0.432 | 0.667 |
| Visit by Site - Vancouver | -0.01087 | 0.02427 | 202.9 | -0.448 | 0.655 |
| Visit<sup>2</sup> by DBS | -0.00014 | 0.00005 | 365.7 | -2.544 | 0.011 |

**Supplementary Table 12:** Regression coefficients for *BDNF* and intellectual enrichment model with cognitive function as outcome variable

| Parameter | Estimate | SE | DF | T-value | P-value |
| --- | --- | --- | --- | --- | --- |
| Intercept | -4.14600 | 0.41450 | 204.8 | -10.003 | <0.001 |
| Visit | 0.31950 | 0.13200 | 549.9 | 2.421 | 0.016 |
| Visit<sup>2</sup> | -0.13430 | 0.03418 | 398.2 | -3.930 | <0.001 |
| DBS | -0.02267 | 0.00259 | 216.8 | -8.764 | <0.001 |
| Age | -0.12860 | 0.01844 | 205.3 | -6.974 | <0.001 |
| Site - London | 1.49100 | 0.49980 | 202.0 | 2.984 | 0.003 |
| Site - Paris | 0.38720 | 0.51440 | 205.8 | 0.753 | 0.452 |
| Site - Vancouver | 0.58970 | 0.50400 | 202.2 | 1.170 | 0.243 |
| Sex - Male | -0.49460 | 0.36980 | 202.3 | -1.337 | 0.183 |
| Antipsychotic medication | -0.96160 | 0.23710 | 686.7 | -4.056 | <0.001 |
| Intellectual Enrichment | 0.36410 | 0.10160 | 202.2 | 3.583 | <0.001 |
| *BDNF* – Met66 allele carriers | -0.21460 | 0.37470 | 202.3 | -0.573 | 0.568 |
| Intellectual Enrichment by *BDNF* | 0.44540 | 0.16140 | 203.5 | 2.760 | 0.006 |
| Visit by age | -0.00492 | 0.00410 | 189.7 | -1.201 | 0.231 |
| Visit by Site - London | -0.23570 | 0.11290 | 192.0 | -2.088 | 0.0381 |
| Visit by Site – Paris | -0.27030 | 0.11450 | 189.0 | -0.236 | 0.814 |
| Visit by Site - Vancouver | -0.21070 | 0.11390 | 196.5 | -1.850 | 0.065 |
| Visit by DBS | -0.00767 | 0.00150 | 494.2 | -5.098 | <0.001 |
| Visit<sup>2</sup> by DBS | 0.00142 | 0.00047 | 397.9 | 2.998 | 0.003 |
| Visit by Intellectual Enrichment | -0.01213 | 0.01836 | 199.9 | -0.661 | 0.510 |
| Visit by *BDNF* | -0.18340 | 0.08423 | 190.5 | -2.177 | 0.031 |

**Supplementary Table 13:** Interaction between intellectual enrichment and *BDNF* on brain volume

| Model | Df | AIC | Chisq | P-value | P-value Bonferroni cor. |
| --- | --- | --- | --- | --- | --- |
| **Caudate volume** |  |  |  |  |  |
| Null model | 21 | -4335.9 |  |  |  |
| + *BDNF* by Intellectual Enrichment | 22 | -4334.1 | 0.2 | 0.654 | 1 |
| + *BDNF* by Intellectual Enrichment by Visit | 23 | -4338.1 | 6.0 | 0.014 | 0.029 |
| **Total GM volume** |  |  |  |  |  |
| Null model | 23 | 645.7 |  |  |  |
| + *BDNF* by Intellectual Enrichment | 24 | 646.9 | 0.8 | 0.376 | 0.751 |
| + *BDNF* by Intellectual Enrichment by Visit | 25 | 637.0 | 11.9 | <0.001 | 0.001 |

The null model included intellectual enrichment, *BDNF*, age at baseline, site (3 dummy variables), DBS at baseline with their interaction with Visit, and sex. For the model with total GM volume as outcome measure a quadratic term was also included for Visit, as well as its interaction with DBS. P-values were corrected for 2 independent comparisons using Bonferroni correction.

**Supplementary Table 14:** Regression coefficients for the *BDNF* and intellectual enrichment model with caudate volume as outcome variable

| Parameter | Estimate | SE | DF | T-value | P-value |
| --- | --- | --- | --- | --- | --- |
| Intercept | -0.01151 | 0.01066 | 203.2 | -1.079 | 0.2818 |
| Visit | 0.01023 | 0.00053 | 187.3 | -19.405 | <0.001 |
| DBS | -0.00067 | 0.00007 | 203.0 | -10.203 | <0.001 |
| Age | 0.00272 | 0.00047 | 203.0 | -5.749 | <0.001 |
| Site - London | 0.02705 | 0.01290 | 203.0 | 2.097 | 0.037 |
| Site - Paris | 0.05544 | 0.01320 | 203.0 | 4.199 | <0.001 |
| Site - Vancouver | -0.00426 | 0.01301 | 203.0 | -0.327 | 0.744 |
| Sex - Male | -0.02385 | 0.00953 | 202.9 | -2.502 | 0.013 |
| Intellectual Enrichment | 0.00143 | 0.00262 | 203.0 | 0.546 | 0.585 |
| *BDNF* – Met66 allele carriers | -0.00541 | 0.00967 | 203.0 | -0.560 | 0.5762 |
| Intellectual Enrichment by *BDNF* | 0.00247 | 0.00416 | 203.0 | 0.595 | 0.553 |
| Visit by Age | 0.00016 | 0.00003 | 188.2 | 6.395 | <0.001 |
| Visit by DBS | -0.00002 | 0.000004 | 186.2 | -6.539 | <0.001 |
| Visit by Site - London | 0.00123 | 0.00068 | 185.6 | 1.801 | 0.073 |
| Visit by Site – Paris | 0.00090 | 0.00071 | 187.8 | 1.272 | 0.205 |
| Visit by Site - Vancouver | -0.00056 | 0.00069 | 191.0 | -0.811 | 0.419 |
| Visit by Intellectual Enrichment | -0.00021 | 0.00014 | 187.7 | -1.518 | 0.131 |
| Visit by *BDNF* | -0.00038 | 0.00052 | 191.8 | -0.727 | 0.468 |
| Visit by Intellectual Enrichment by *BDNF* | 0.00057 | 0.00023 | 203.8 | 2.466 | 0.015 |

**Supplementary Table 15:** Regression coefficients for the *BDNF* and intellectual enrichment model with total GM volume as outcome variable

| Parameter | Estimate | SE | DF | T-value | P-value |
| --- | --- | --- | --- | --- | --- |
| Intercept | 1.12900 | 0.47320 | 207.1 | 2.387 | 0.018 |
| Visit | -0.07176 | 0.02103 | 332.0 | -3.412 | 0.001 |
| Visit<sup>2</sup> | -0.03101 | 0.00406 | 347.5 | -7.645 | <0.001 |
| DBS | -0.01205 | 0.00298 | 197.2 | -4.042 | <0.001 |
| Age | -0.21060 | 0.02171 | 198.2 | -9.700 | <0.001 |
| Site - London | -0.69290 | 0.58020 | 197.1 | -1.194 | 0.234 |
| Site - Paris | -0.64750 | 0.59370 | 197.3 | -1.091 | 0.277 |
| Site - Vancouver | -0.10070 | 0.59790 | 198.8 | -0.168 | 0.866 |
| Sex - Male | -1.48000 | 0.37870 | 194.5 | -3.909 | <0.001 |
| Intellectual Enrichment | -0.04635 | 0.11830 | 199.2 | -0.392 | 0.696 |
| *BDNF* – Met66 allele carriers | -0.26430 | 0.43790 | 198.3 | -0.603 | 0.547 |
| Intellectual Enrichment by *BDNF* | 0.16480 | 0.18900 | 197.8 | 0.872 | 0.384 |
| Visit by Age | -0.00460 | 0.00088 | 186.9 | -5.204 | <0.001 |
| Visit by DBS | -0.00042 | 0.00020 | 503.9 | -2.075 | 0.039 |
| Visit by Site - London | 0.01646 | 0.02304 | 183.8 | 0.714 | 0.476 |
| Visit by Site – Paris | -0.00322 | 0.02381 | 187.2 | -0.135 | 0.893 |
| Visit by Site - Vancouver | -0.00675 | 0.02426 | 190.5 | -0.278 | 0.781 |
| Visit by Intellectual Enrichment | -0.01016 | 0.00459 | 178.6 | -2.212 | 0.028261 |
| Visit by *BDNF* | -0.01294 | 0.01752 | 187.0 | -0.738 | 0.461 |
| Visit by Intellectual Enrichment by *BDNF* | 0.02680 | 0.00763 | 193.0 | 3.511 | <0.001 |
| Visit<sup>2</sup> by DBS | -0.00015 | 0.00006 | 347.7 | -2.701 | 0.007253 |

**Supplementary Table 16:** Association between control risk factors and cognitive function

| Model | Df | AIC | Chisq | P-value |
| --- | --- | --- | --- | --- |
| **Antipsychotic medication** |  |  |  |  |
| Null model | 19 | 3328.3 |  |  |
| + Antipsychotic medication | 20 | 3316.2 | 14.1 | <0.001 |
| + Antipsychotic medication by Visit | 21 | 3316.6 | 1.6 | 0.200 |
| **Depression** |  |  |  |  |
| Null model | 19 | 3242.3 |  |  |
| + Depression | 20 | 3240.6 | 3.8 | 0.052 |
| + Depression by Visit | 21 | 3242.5 | 0.0 | 0.913 |
| **ApoE e3 allele carriers** |  |  |  |  |
| Null model | 19 | 3046.3 |  |  |
| + Intellectual Enrichment | 20 | 3048.3 | 0.1 | 0.794 |
| + Intellectual Enrichment by Visit | 21 | 3047.4 | 2.8 | 0.092 |

The null model included age at baseline, DBS at baseline, site (3 dummy variables) with their interaction with Visit, and sex. A quadratic term was also included for Visit, as well as its interaction with DBS.

## Supplementary Figures

**Supplementary Figure 1:**
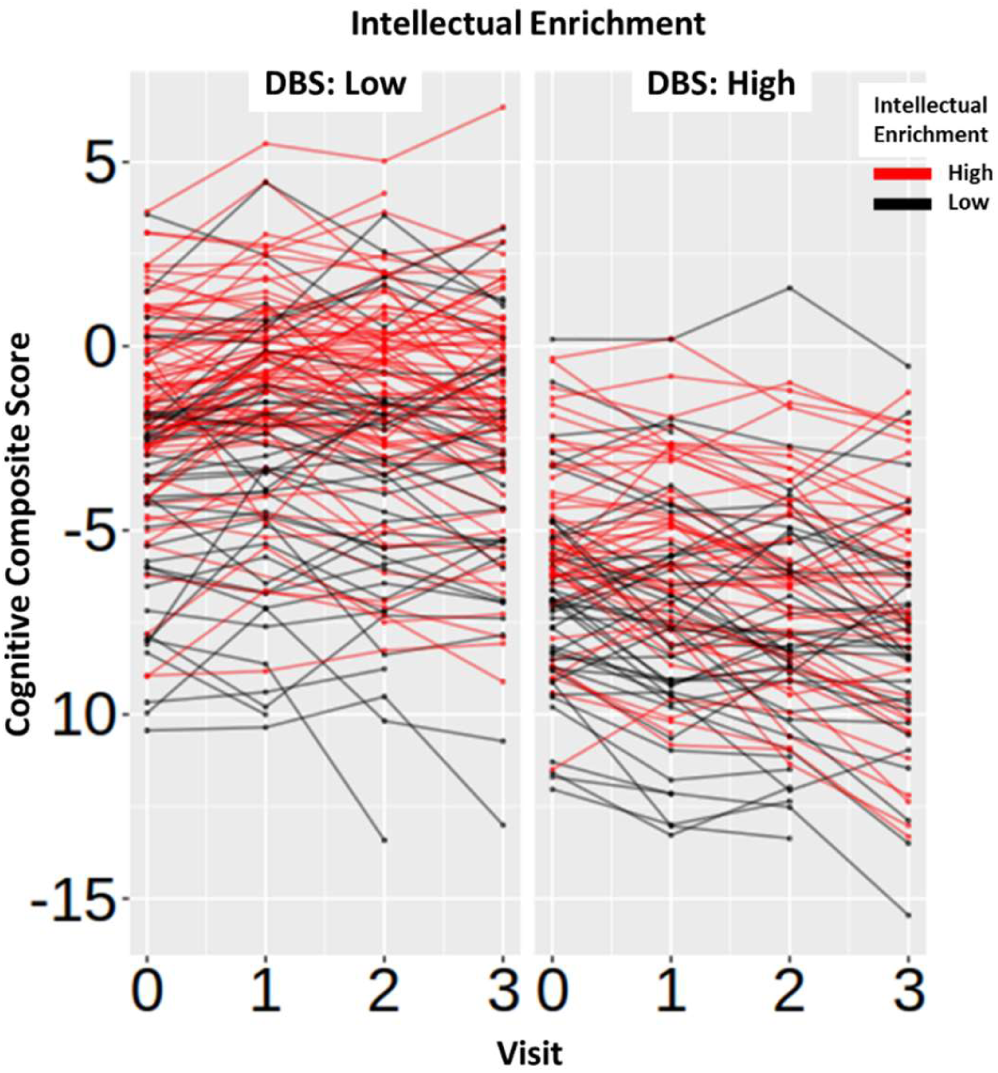
Association between intellectual enrichment, DBS and global cognitive function. For visualization purposes results are split into high (above mean) and low (below mean) DBS. Individual lines are drawn for each participant and colour coded for high (above mean; red) and low (below mean; black) intellectual enrichment. Datapoints show the raw data residualized against age, site, sex and use of antipsychotic medication.

**Supplementary Figure 2:**
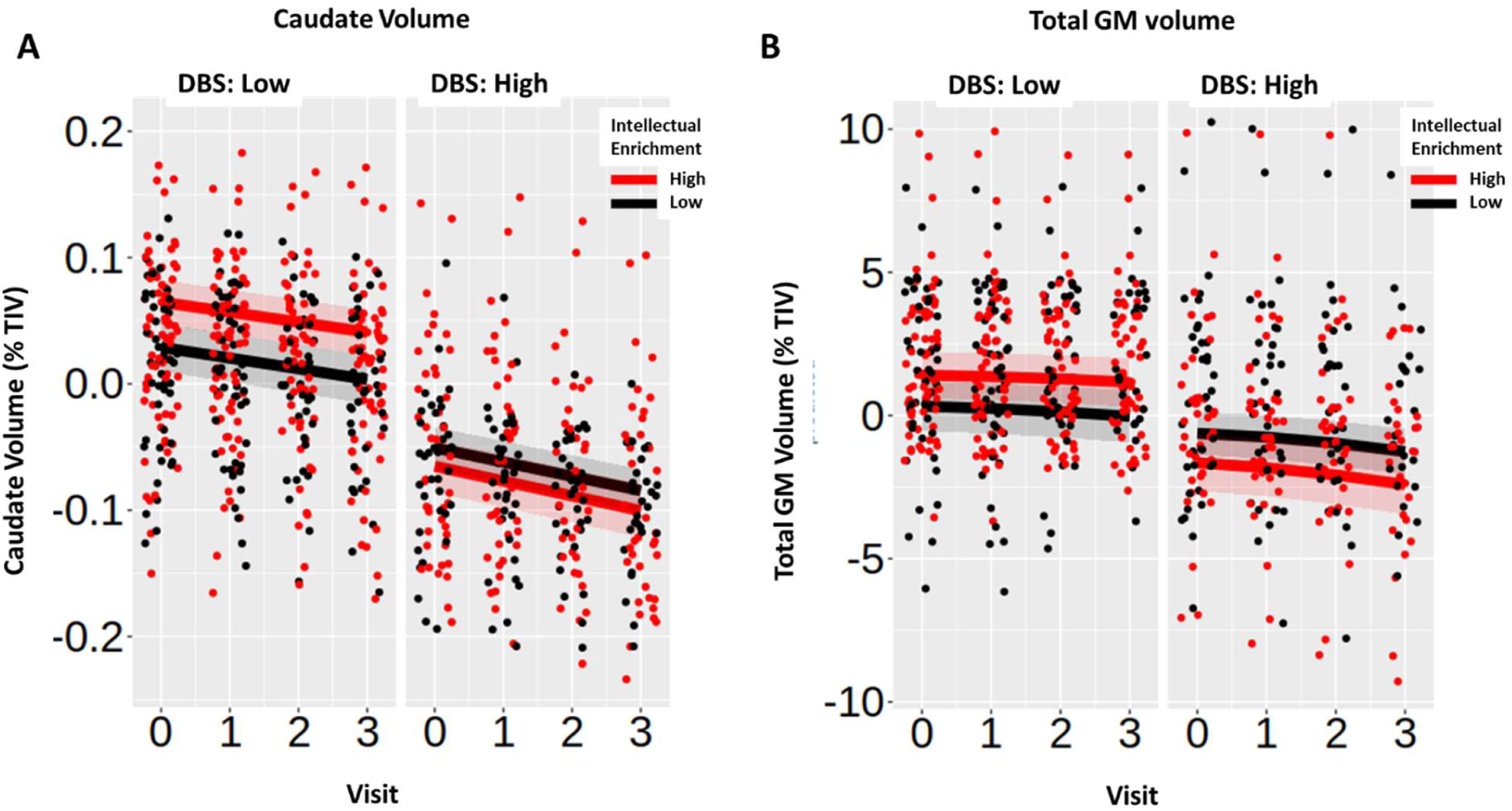
Association between intellectual enrichment and DBS with caudate volume (A) and total GM volume (B) as percent of total intracranial volume (TIV). For visualization purposes results are split into high (above mean) and low (below mean) DBS. Regression lines are generated from the mixed linear model at high (1SD above mean; red) and low (1SD below mean; black) intellectual enrichment. Bands around the regression lines are 95% confidence intervals. Datapoints show the raw data residualized against age, site, sex and use of antipsychotic medication, and have been jittered to minimize overlap.

**Supplementary Figure 3:**
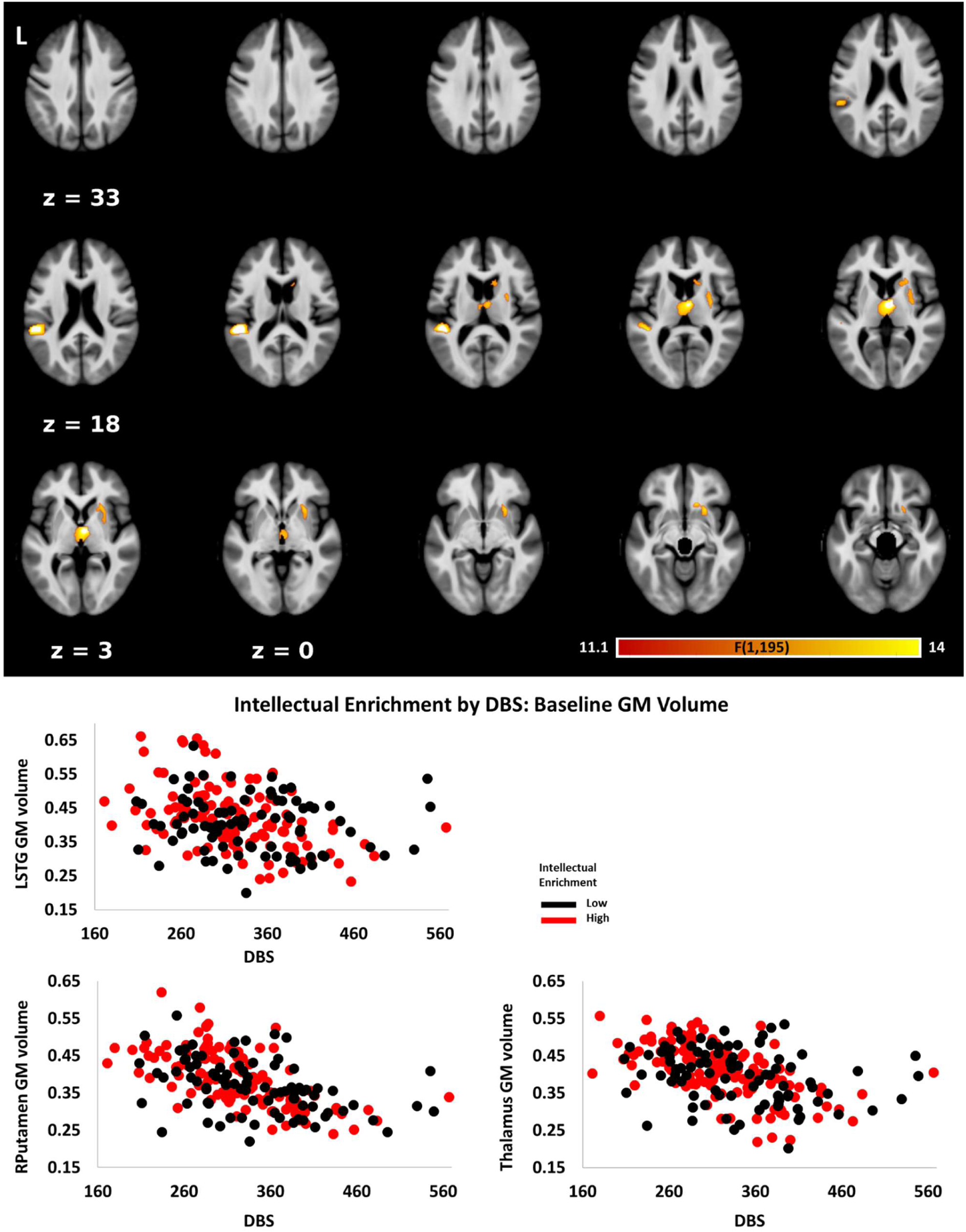
Association between intellectual enrichment and DBS with baseline GM volume. Shown on top are significant clusters overlaid on the group template. Maps are thresholded at p < 0.001 uncorrected at voxel level and p < 0.05 family-wise error (FWE) corrected at cluster-level. Shown in a scatter plots (bottom) are the extracted values averaged across the 3 significant clusters. For visualization purposes data are grouped by high (above mean - red) and low (below mean - black) intellectually enrichment. Data have been adjusted for age, site, sex and TIV. LSTG = left superior temporal gyrus.

**Supplementary Figure 4:**
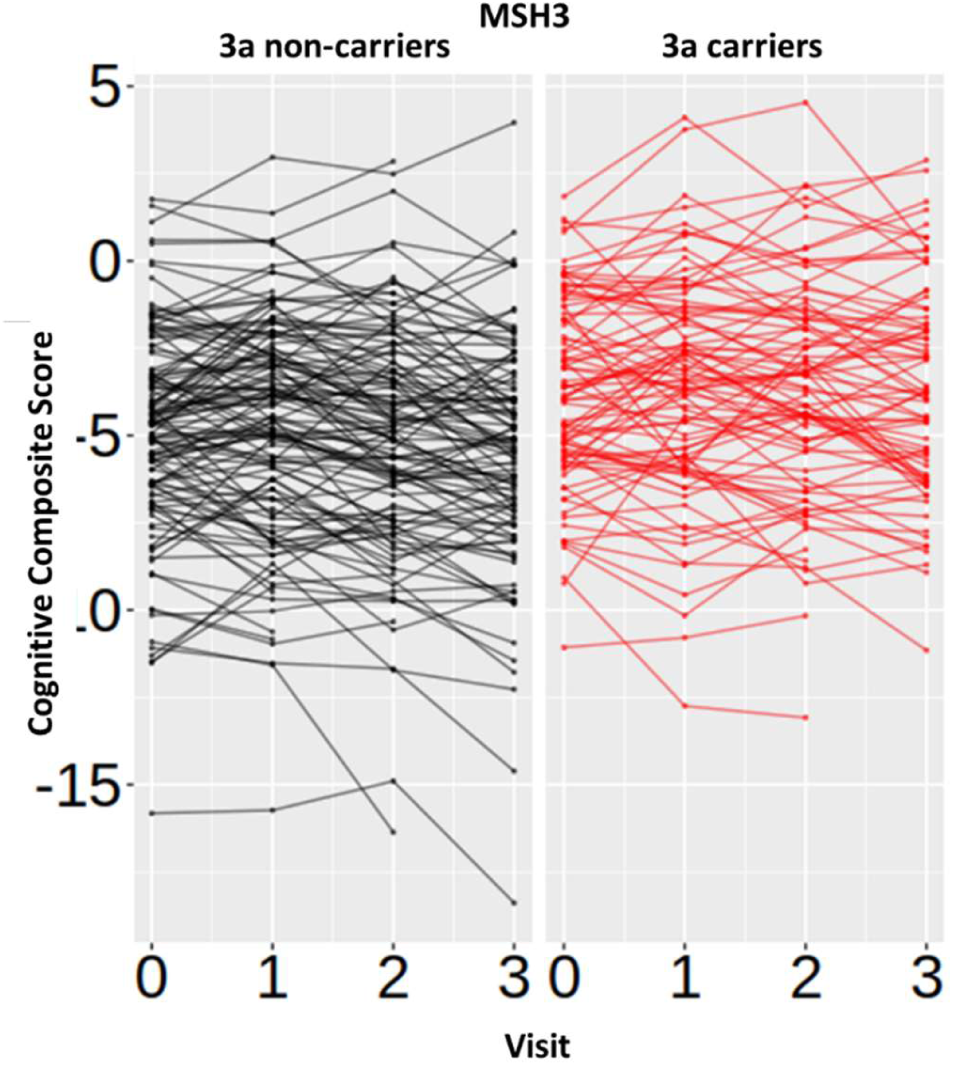
Association between the MSH3 predictor with global cognitive function. For visualization purposes results are shown in separate panels for carriers (right, red) and non-carriers (left, black) of the 3a allele. Individual lines are drawn for each participant. Datapoints show the raw data residualized against age, DBS, site, sex and use of antipsychotic medication.

**Supplementary Figure 5:**
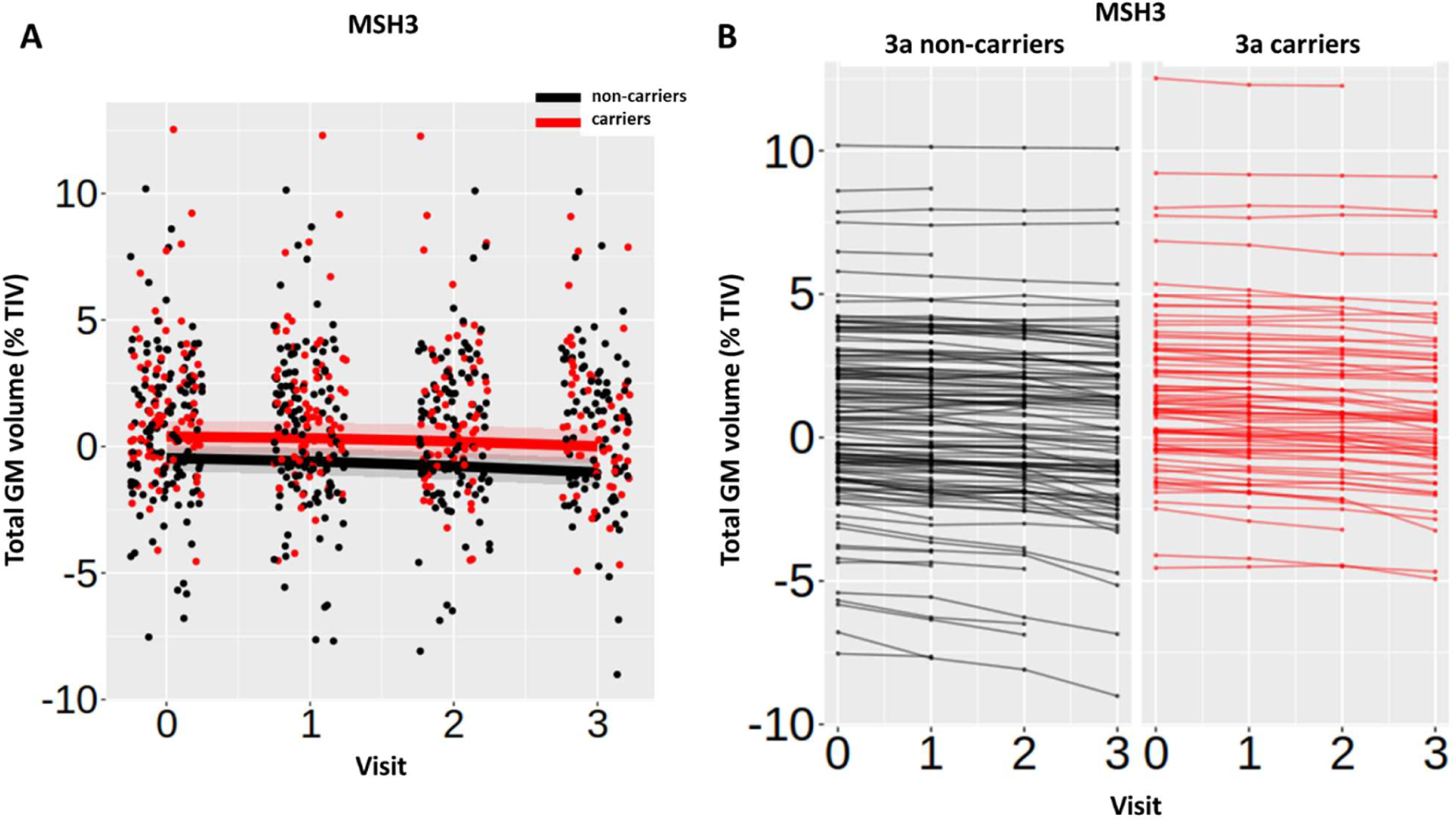
Association between the MSH3 predictor with total GM volume as percent TIV. In (A) regression lines are generated from the mixed linear model for carriers (red) and non-carriers (black) of the 3a allele. Bands around the regression lines are 95% confidence intervals. Datapoints have been jittered to minimize overlap. In (B) individual lines are drawn for each participant and colour coded for carriers (right, red) and non-carriers (left, black) of the 3a allele. Datapoints show the raw data residualized against age, DBS, site and sex.

**Supplementary Figure 6:**
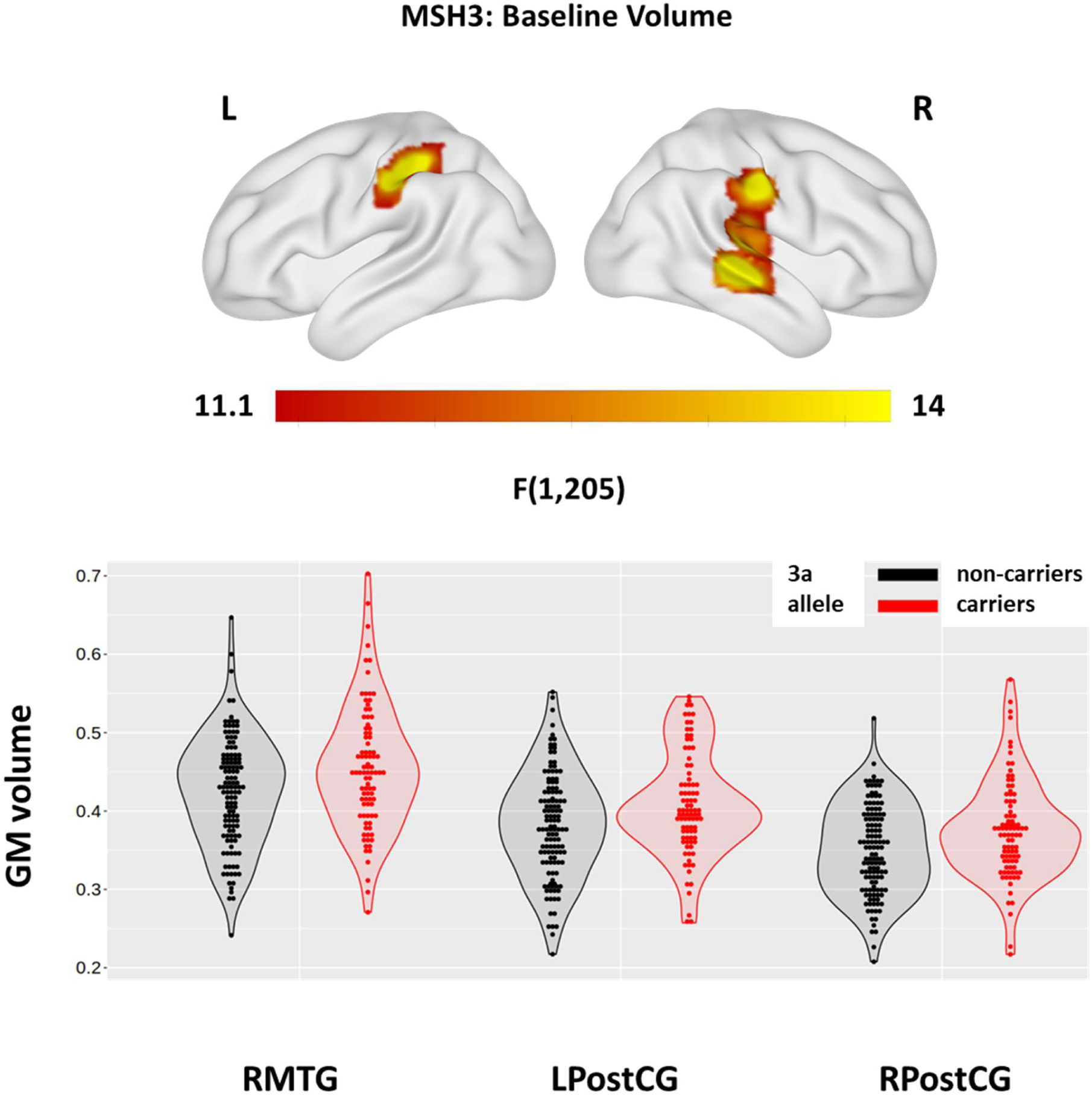
Association between the *MSH3* predictor with baseline GM volume. Significant clusters are overlaid on the ICBM152 template mesh (top). Maps are thresholded at p < 0.001 uncorrected at voxel level and p < 0.05 family-wise error (FWE) corrected at cluster-level. Shown in violin plots (bottom) are the extracted values averaged across the 3 significant clusters for carriers (red) and non-carriers (black) of the 3a allele. Individual datapoints are shown in black dots. Datapoints show the raw data residualized against age, DBS, site, sex and TIV. RMTG = right middle temporal gyrus; LPostCG and RPostCG= left and right postcentral gyrus.

**Supplementary Figure 7:**
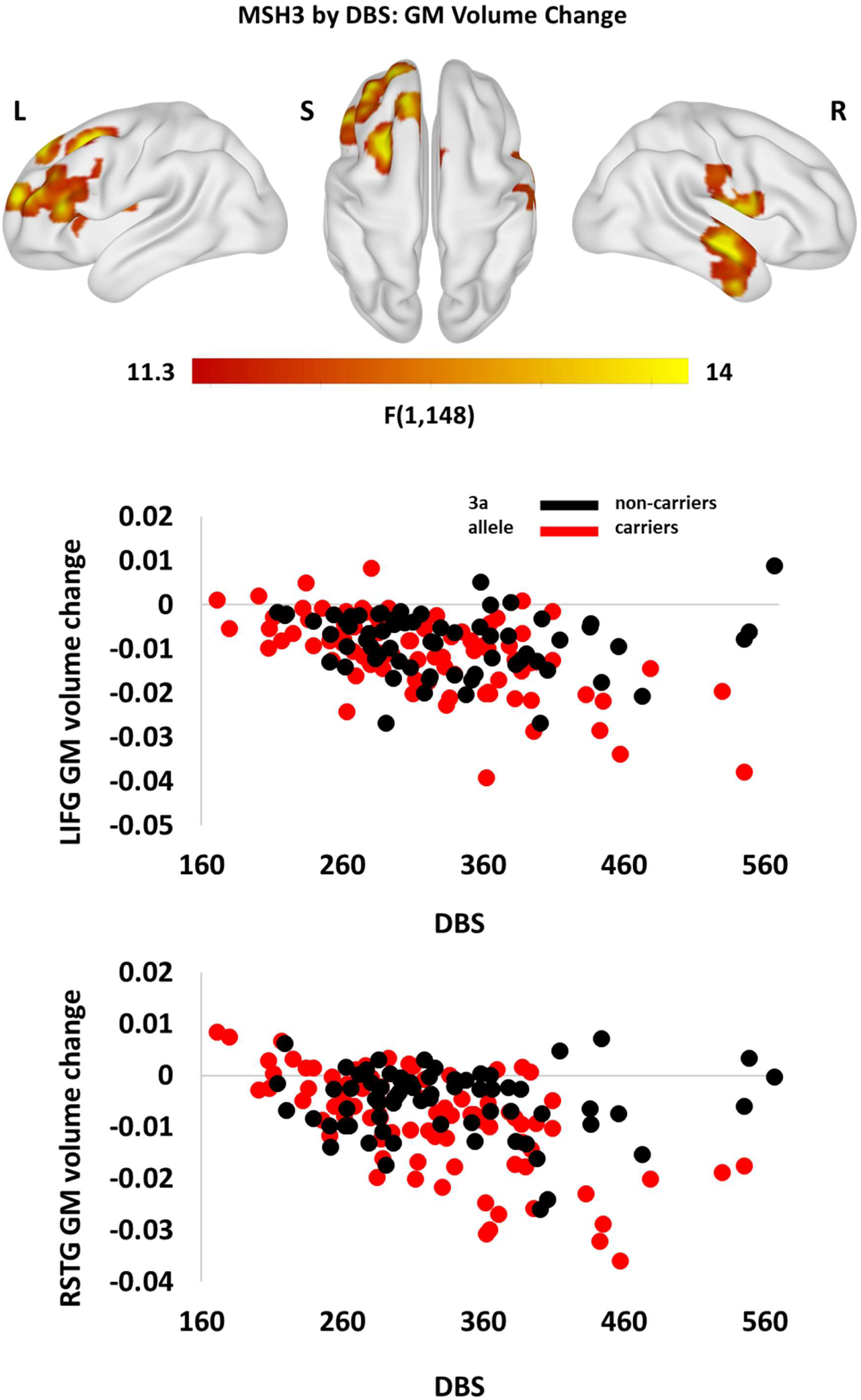
Association between the *MSH3* predictor and DBS with 3-year GM volume change (B). Significant clusters are overlaid on the ICBM152 template mesh (top). Maps are thresholded at p < 0.001 uncorrected at voxel level and p < 0.05 family-wise error (FWE) corrected at cluster-level. Shown in scatter plots (bottom) are the extracted values averaged across the significant clusters for carriers (red) and non-carriers (black) of the 3a allele. Datapoints show the raw data residualized against age, DBS, site, sex and TIV. RSTG = right superior temporal gyrus; LIFG = left and right inferior frontal gyrus.

**Supplementary Figure 8:**
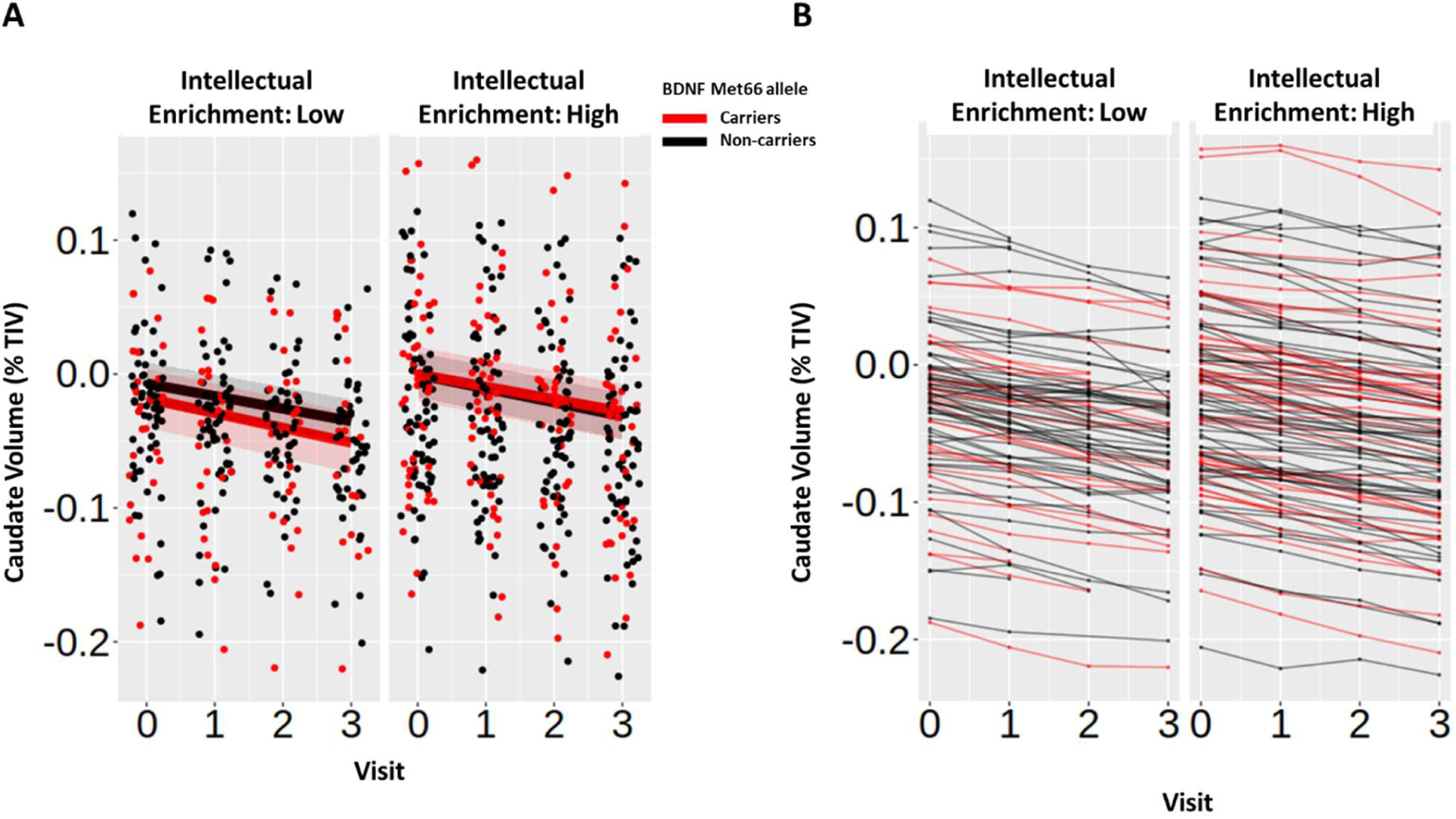
Association between intellectual enrichment and *BDNF* with caudate volume as percent TIV. For visualization purposes results are split into high (above mean) and low (below mean) intellectual enrichment. In (A) regression lines are generated from the mixed linear model for *BDNF* Met66 allele carriers (red) and non-carriers (black). Bands around the regression lines are 95% confidence intervals. Data have been jittered to minimize overlap. In (B) individual lines are drawn for each participant and colour coded for carriers (red) and non-carriers (black) of the Met66 allele. Datapoints in both plots show the raw data residualized against age, DBS, site and sex.

**Supplementary Figure 9:**
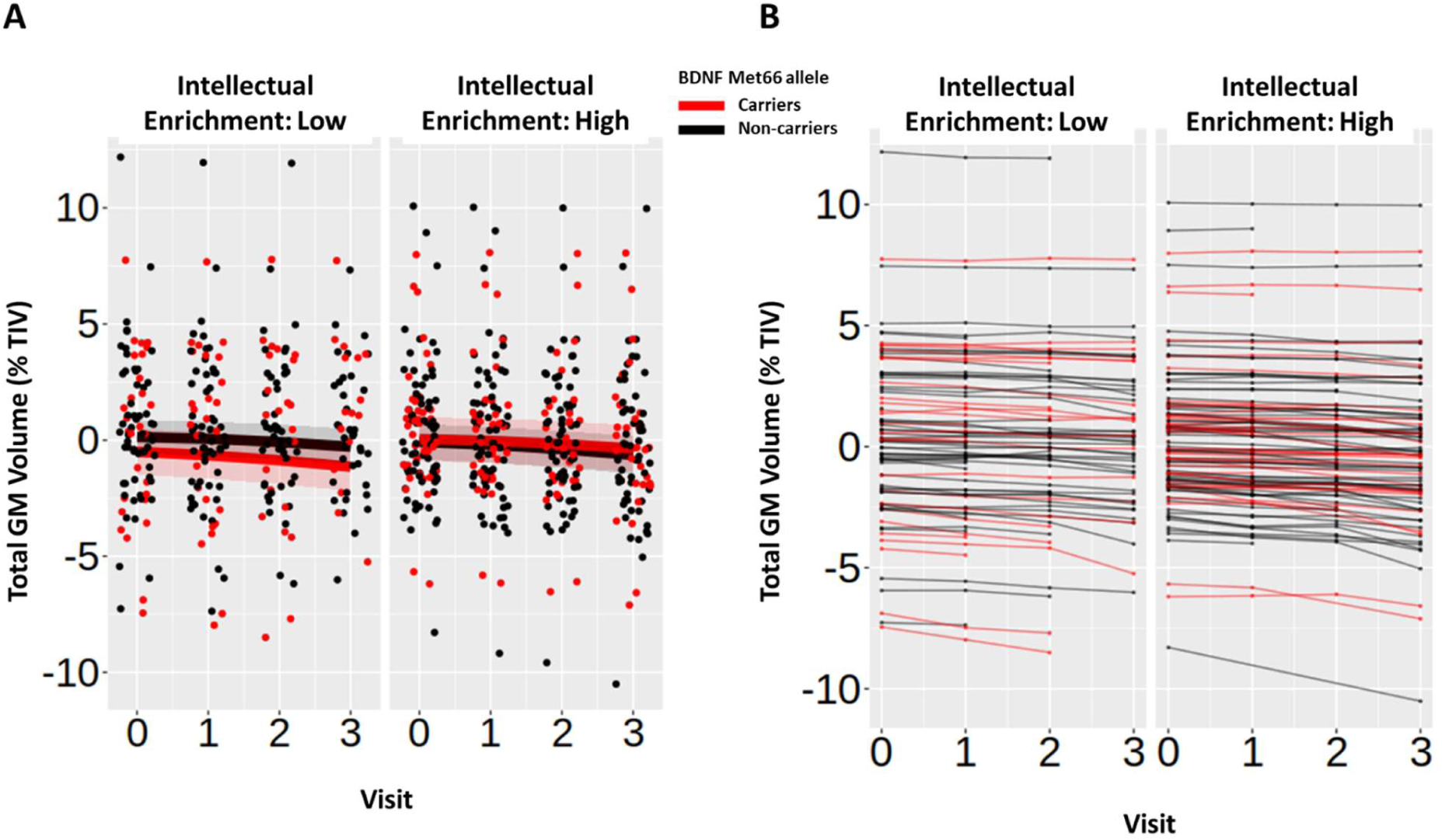
Association between intellectual enrichment and *BDNF* with total GM volume as percent TIV. For visualization purposes results are split into high (above mean) and low (below mean) intellectual enrichment. In (A) regression lines are generated from the mixed linear model for *BDNF* Met66 allele carriers (red) and non-carriers (black). Bands around the regression lines are 95% confidence intervals. Data have been jittered to minimize overlap. In (B) individual lines are drawn for each participant and colour coded for carriers (red) and non-carriers (black) of the Met66 allele. Datapoints in both plots show the raw data residualized against age, DBS, site and sex.

